# Evaluating Precision and Responsiveness of MRI-derived Brain Volumetric and Clinical Outcome Measures in CLN3 Disease

**DOI:** 10.64898/2026.09.29.26363931

**Authors:** Rakibul Hafiz, Cristan Farmer, Lisa Joseph, Ariane Soldatos, Amritha Nayak, Audrey E Thurm, An N. Dang Do, Carlo Pierpaoli

## Abstract

**Background and Objective:** For their acceptance as endpoints in clinical trials, biomarkers and clinical outcome assessment (COA) measures should show reliability by being reproducible (precise) and responsive to change. We evaluated precision and responsiveness of MRI-derived brain volume metrics and various COAs in ceroid-lipofuscinosis, neuronal-3 (CLN3) disease.

**Methods:** This longitudinal observational study (NCT03307304) included 44 individuals with CLN3-related disease (ages 0.5–29.1 years, 48% female). Brain volumetric measures were computed using SynthSeg-v2.0 from T1-weighted 3D MPRAGE images acquired at 3T (Median [Q1, Q3] total MRI follow-up duration from baseline: 2.04 [0.00, 3.69] years). Compartmental tissue volumes were normalized to a model-adjusted total intracranial volume and scaled to the tissue-specific volume fraction expected in healthy adolescent brains to extract measures of brain atrophy. COAs included the Vineland Adaptive Behavior Scales, verbal IQ, and the Unified Batten Disease Rating Scale (UBDRS). Analysis of linear mixed-effects models included only CLN3 participants with syndromic presentation, having three or more visits per measurement to estimate slope, its relative standard error (RSE), and the minimum inter-visit interval (*T_min_*) for statistically significant progression for each participant. *T_min_* to detect progression in 75% of the population (*T_min,75%_*) was computed for each metric.

**Results:** Among brain volumetric measures, brain parenchyma volume (BPV) and gray matter volume (GMV) were the most precise and responsive metrics. BPV showed an overall *intra*-individual decline with age of −1.32%/year (95% CI: −1.64% to −0.99%), low RSE of 12.5%, and short *T_min,75%_* of 0.85 years (95% CI: 0.47–1.24). Amongst COAs, the UBDRS Clinical Global Impression (CGI) score was the most precise and responsive with an overall *intra*-individual increase of 1.26 units/year (95% CI: 1.03–1.49), RSE of 24.10%, and *T_min,75%_* of 2.56 years (95% CI: 1.73–3.46), though with lower precision (RSE higher) and lower responsiveness (*T_min,75%_* longer) than BPV.

**Discussion:** MRI-derived metrics, in particular BPV and GMV, detected change in brain atrophy with high precision and responsiveness suggesting that they would be informative for evaluating potential neuroprotective therapies in CLN3 disease. These findings support incorporating MRI-derived volumetric measurements alongside clinical assessments in future clinical trials in CLN3.

## INTRODUCTION

Ceroid-lipofuscinosis, neuronal 3 (CLN3) is the most common type of neuronal ceroid lipofuscinosis (NCL) in pediatric cohorts, with onset typically in early childhood.^1–3^ It is a rare, autosomal recessive, neurodegenerative disease that typically presents with progressive vision loss, followed by declining cognitive and psycho-motor function and seizures. CLN3 is grouped with other lysosomal diseases based on intra-lysosomal accumulation of auto-fluorescent, fatty-proteinaceous material (ceroid lipofuscin).^1^

CLN3 disease is usually fatal by early to mid-adulthood^4^, and currently has no cure. Experimental therapies hypothesized to slow the progression (e.g., miglustat^5, 6^, gemfibrozil^7^) or correct the underlying defect (e.g., gene therapy^8, 9^, antisense oligonucleotides^10, 11^) have been tested only in animals or small numbers of individuals. Trial outcome measures and substantiated approaches for individualized treatment evaluation continue to be investigated. The clinician-rated Unified Batten Disease Rating Scale^12, 13^ (UBDRS) tracks with age and disease state.

Neurocognitive performance-based outcome measures (e.g., verbal IQ from the Wechsler scales) and caregiver-reported adaptive behavior (e.g., Vineland Adaptive Behavior Scales) assessments have shown strong correlations with age and disease state in cross-sectional and limited longitudinal data.^14, 15^ Although these COAs measure meaningful aspects of health, and are therefore necessary to be included as endpoints in trials, their dependence on individual performance and caregiver report impacts their reliability. Quantitative, objective measures that correlate with COAs would strengthen the toolbox for evaluating CLN3 disease.

Neuroimaging studies of CLN3 disease have revealed brain atrophy globally in gray and white matter^16^ and, regionally, in the hippocampus.^17^ Here we investigate the precision and responsiveness of T1-weighted (T1w) MRI-derived brain compartment volume measurements, with particular focus on brain parenchyma volume (BPV), as candidate biomarkers of atrophy and disease progression. We apply the same analytic framework to several clinical outcome assessments (COAs) widely used in CLN3.

This study focuses on tracking progression and finding potential endpoints to assess efficacy of experimental therapy. To be used as a clinical trial endpoint, investigators must provide sufficient evidence that COAs measure and biomarkers predict meaningful aspects of health. Equally important, however, is evidence indicating that the tool’s measurement is reliable and, within its intended context of use, responsive to change. This aspect is comparatively neglected in the CLN3 literature, for both biomarkers and COA metrics. Precision and responsiveness are nonetheless of particular importance for rare diseases such as CLN3, where the number of individuals available for any given trial is necessarily small, and where a measure’s ability to reliably detect a change at the individual level, within a feasible observation window, directly determines statistical power. A measure with high individual-level noise or a long minimum interval to detect statistically meaningful change may correlate well with disease state on average across a cohort– yet still be poorly suited to a trial that must rely on a small number of participants and a limited follow-up duration. Leveraging the concurrently collected imaging and clinical data in this study, we evaluate the precision and responsiveness of MRI-derived BPV alongside several commonly used COAs to address two fundamental questions:

i. if a given measurement changes with age due to disease progression, how precisely can this within-person rate of change be estimated? and
ii. what is the minimum inter-visit interval required to detect a within-individual statistically significant change in the direction of disease progression. Furthermore, what is the minimum duration for a large enough proportion (e.g., 75%) of the population to reach a significance to reliably document CLN3 progression?

## METHODS

### CLN3 Participant Demographics

This is a prospective, longitudinal observational study of individuals with genetic, laboratory, or clinical evidence of CLN3-related disease (NCT03307304), conducted at the NIH Clinical Center. Recruitment and evaluation were approved by the NIH Institutional Review Board and performed per International Conference on Harmonization Good Clinical Practice guidelines and applicable US Code of Federal Regulations. Written informed consent, and assent where appropriate, was obtained from all participants. Follow-up occurs annually as feasible.

This analysis included participants enrolled and imaged through May 15, 2026. As of the cutoff date, the sample comprised 44 individuals based on our inclusion criteria for the baseline age to be < 35 years (baseline age range: 0.5–29.1 years; excluding SP.33.2.2 and SP42.2.2) and baseline intervention status to be pre-treatment (excluding SP.44.2.2). On average – each individual had a repeated MRI scan at ∼1-year intervals (median [Q1, Q3] inter-visit duration = 1.02 [1.00, 1.11] years). Overall, the participants had a total follow-up duration from the baseline visit of 2.04 [0, 3.69] years. This cohort include three participants <5 years of age at enrolment (‘Early-Childhood’– two pre-symptomatic: no disease-related changes and one with only vision changes), as well as three >13 years of age at enrolment with only vision changes (‘Vision-Only’: no decline in cognitive or psychomotor function). For more details on individual participant demographics, please refer to eTable 1.

The early-childhood and vision-only subgroups differed meaningfully from the rest of the cohort on visual inspection: the early-childhood participants (SP19.2.1, SP39.2.2 and SP43.2.4) showed typical brain development, and vision-only participants (SP5.2.1, SP5.2.2 and SP29.2.2) had remarkably less atrophic/stable longitudinal BPVs. Including them would introduce heterogeneity and skewness into an otherwise homogeneous sample, violating key assumptions of the statistical models used. Due to their small number, subgroup analysis was not possible. We therefore retained both subgroups for descriptive and visualization purposes only. Data obtained from participants/participant visits on acetylleucine, miglustat, gemfibrozil, or gene replacement therapies were excluded. Formal statistical analyses were restricted to 38 participants (50% female; 7 sibling pairs) with syndromic CLN3 presentation, comprising 107 total MRI observations (baseline age range: 6.0–20.7 years). MRI visits per participant ranged from one to six. Baseline characteristics of the syndromic CLN3 cohort are summarized in Table 1; demographic composition by MRI visit and age is shown in eFigure 1 (eAppendix).

**Figure 1.**
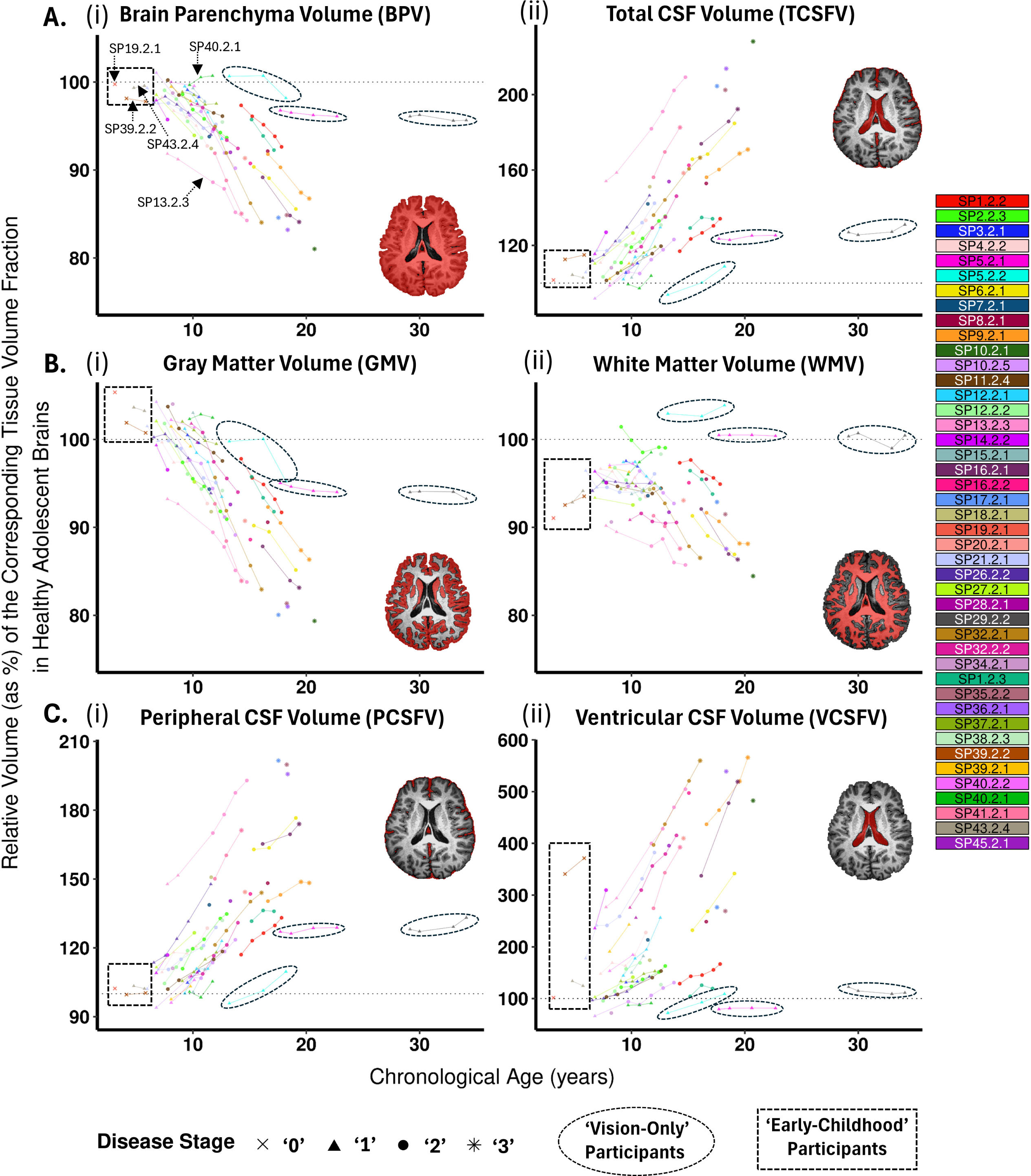
Longitudinal natural history trends in CLN3 individuals for volumetric measurements. The ‘Vision-Only’ participants (dashed ellipses) can be easily distinguished from the rest of the cohort in all panels. The single data points with no linear trends are for participants with a single visit up to the cut-off date. This figure showcases the volumetric trends from the entire sample of N = 44 participants. The shape of the data points represents the disease stage of individuals across age. Most asterisks (* = Stage ‘3’) appear around the age of 20 and beyond, indicating high disease severity at later ages. The dashed boxes identify the ‘Early-Childhood’ participants who show characteristic development (relatively stable BPV values, as opposed to atrophy) at very young ages (< 5 years). The brain map in each panel shows a segmentation binary mask for each corresponding brain compartment. The voxels in red show the typical spatial extent over which the volume is computed for each tissue class. The brain parenchyma and gray matter volumes (**A**(i): BPV and **B**(i): GMV, respectively) show highly consistent downward trends with very low variability in measurements. Atrophy in the white matter (**B**(ii): WMV) is comparatively less (majorly in gray matter) and the measurements are relatively more variable. The compensatory increase in total CSF volume (TCSFV) due to loss in brain parenchyma can be observed in **A**(ii), which is also reflected in the constituent peripheral and ventricular CSF volumes (**C**(i): PCSFV and **C**(ii): VCSFV, respectively). Therefore, the volumetric measurements reliably capture progressive neurodegeneration across the natural history of CLN3 individuals. Note, some participant IDs are labeled in **A**(i): two of them (SP13.2.3 and SP40.2.1) are specifically labeled for a more personalized assessment of exceptional cases. Three additional IDs (SP19.2.1, SP39.2.2 and SP43.2.4) are also labeled; to identify the ‘Early-Childhood’ participants bound within the dashed boxes in all panels. Each participant is identified by a unique color shown in the color-bar (on the right) and these attributes are shared across all metrics. *Keys:* CSF = Cerebrospinal Fluid.

**Table 1.**
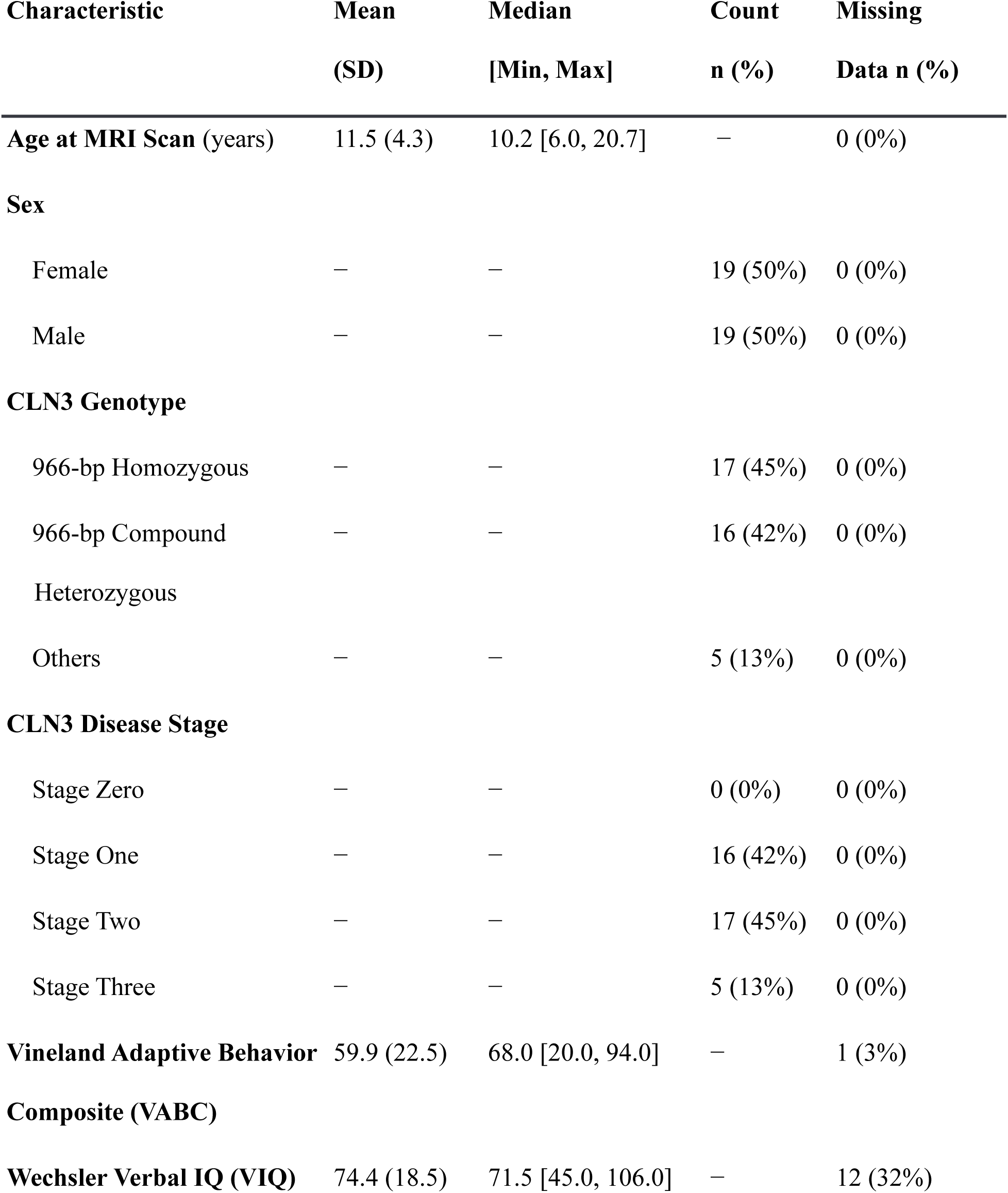
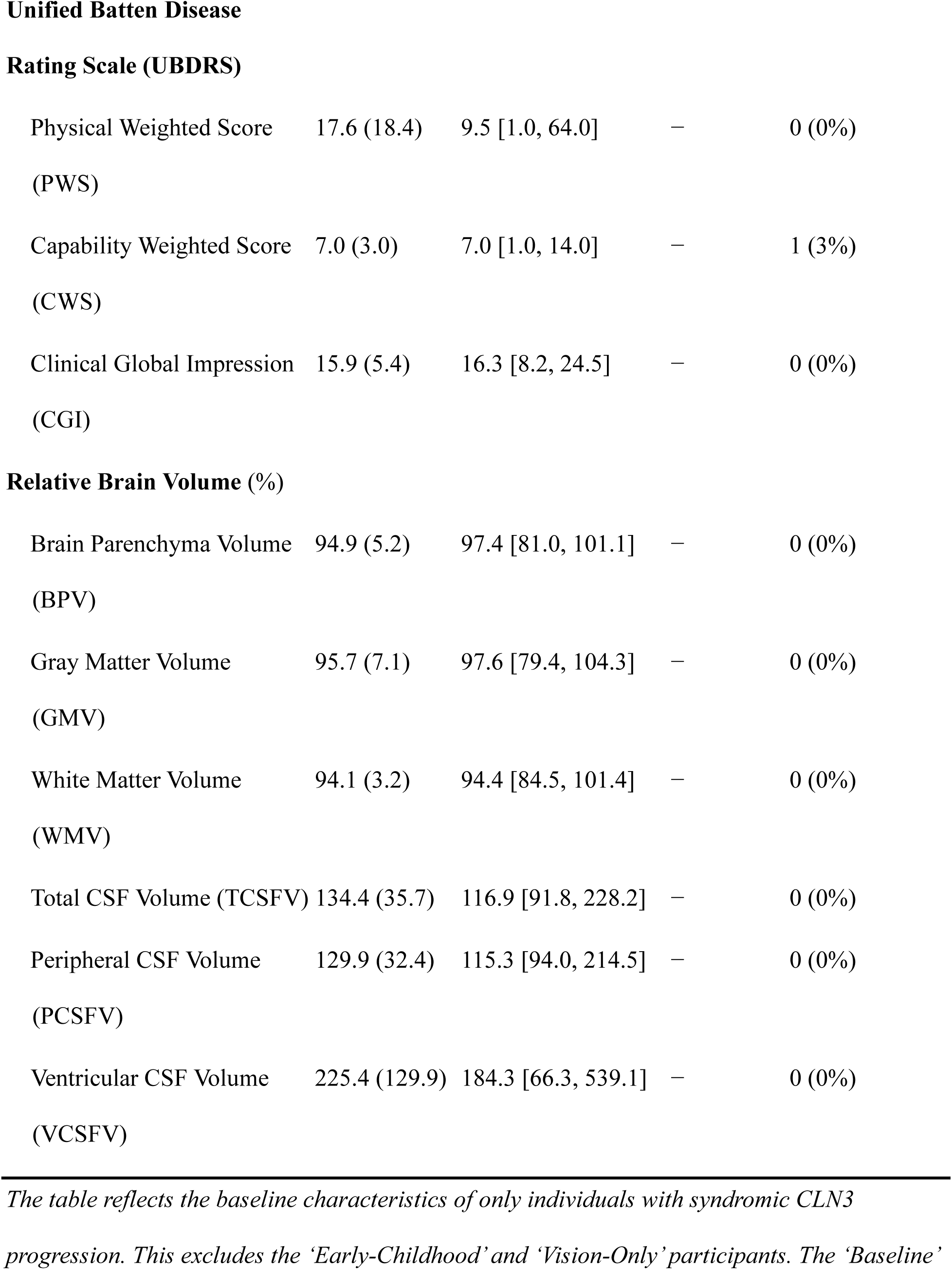
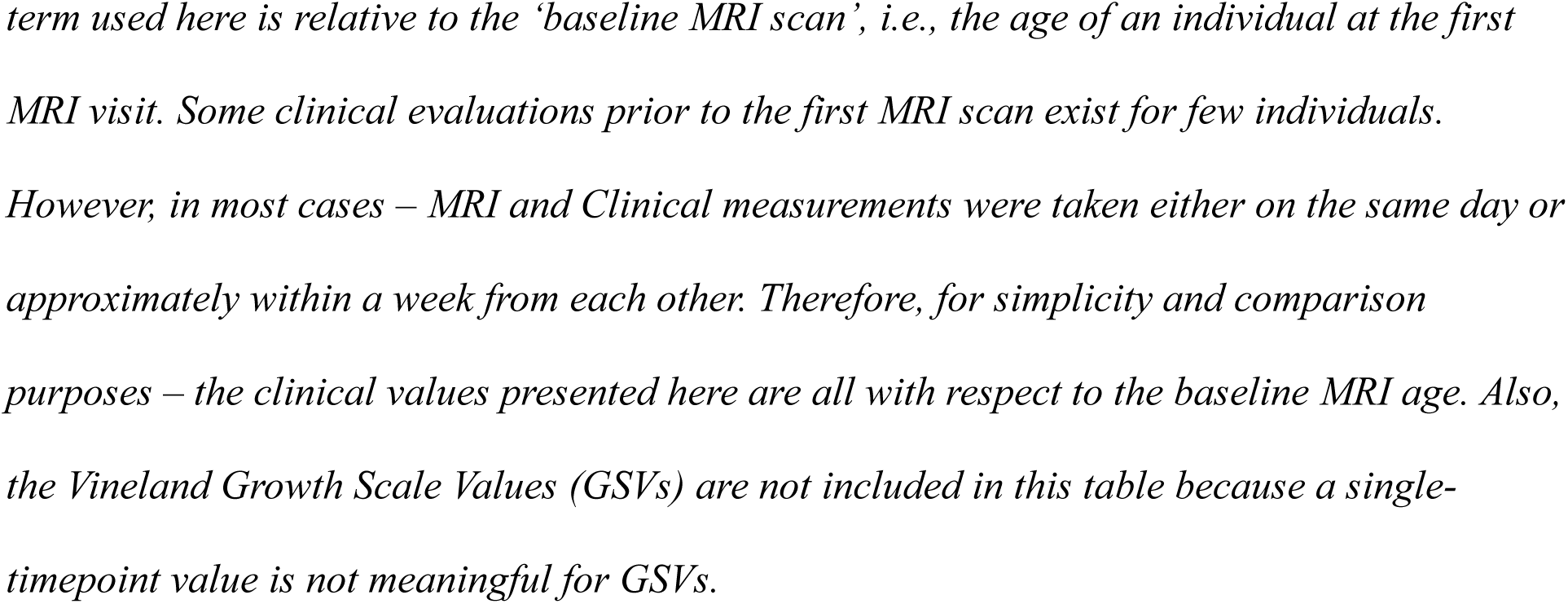
Baseline Characteristics for Syndromic CLN3 Participants (N=38)

MRI was acquired under sedation, and clinical assessments were obtained concurrently, or in close proximity (within the week, if possible) to MRI acquisition visits. In addition, clinical assessments were also obtained from participants who could not travel to NIH (e.g., because of COVID-19 travel restrictions or advanced disease state) and thus could not undergo imaging. As a result, the subset of the 38 syndromic CLN3 participants with sufficient repeated measurements for slope-precision analyses (see Statistical Analysis) differs in size across measurements.

### T1-weighted MRI Imaging

All participants were scanned under propofol sedation on a 3T Philips Achieva scanner using a T1-weighted 3D MPRAGE sequence (TR = 10.586 ms, TE = 5.104 ms, flip angle = 8°, FOV = 240 × 240 mm, resolution = 0.94 × 0.94 × 1.00 mm³, total acquisition time = 8 minutes 30 seconds).

### Quality Control Assessment

All T1w images were visually inspected for imaging and motion artifacts (minimal given sedation). One visit’s T1w-data was excluded due to severe artifacts. All segmentation maps were overlaid on their corresponding T1w image and inspected for segmentation accuracy.

### Volumetric Measurements

There are several novel, informative and technical aspects in the pipeline used to compute the brain compartment volumes to accurately measure brain atrophy in this study. The segmentation methodology and the normalization of raw brain volumes to generate a direct measure for atrophy are detailed in the eAppendix (see eMethods: Sections 1.1 – 1.3 and eFigures 2 and 3).

**Figure 2.**
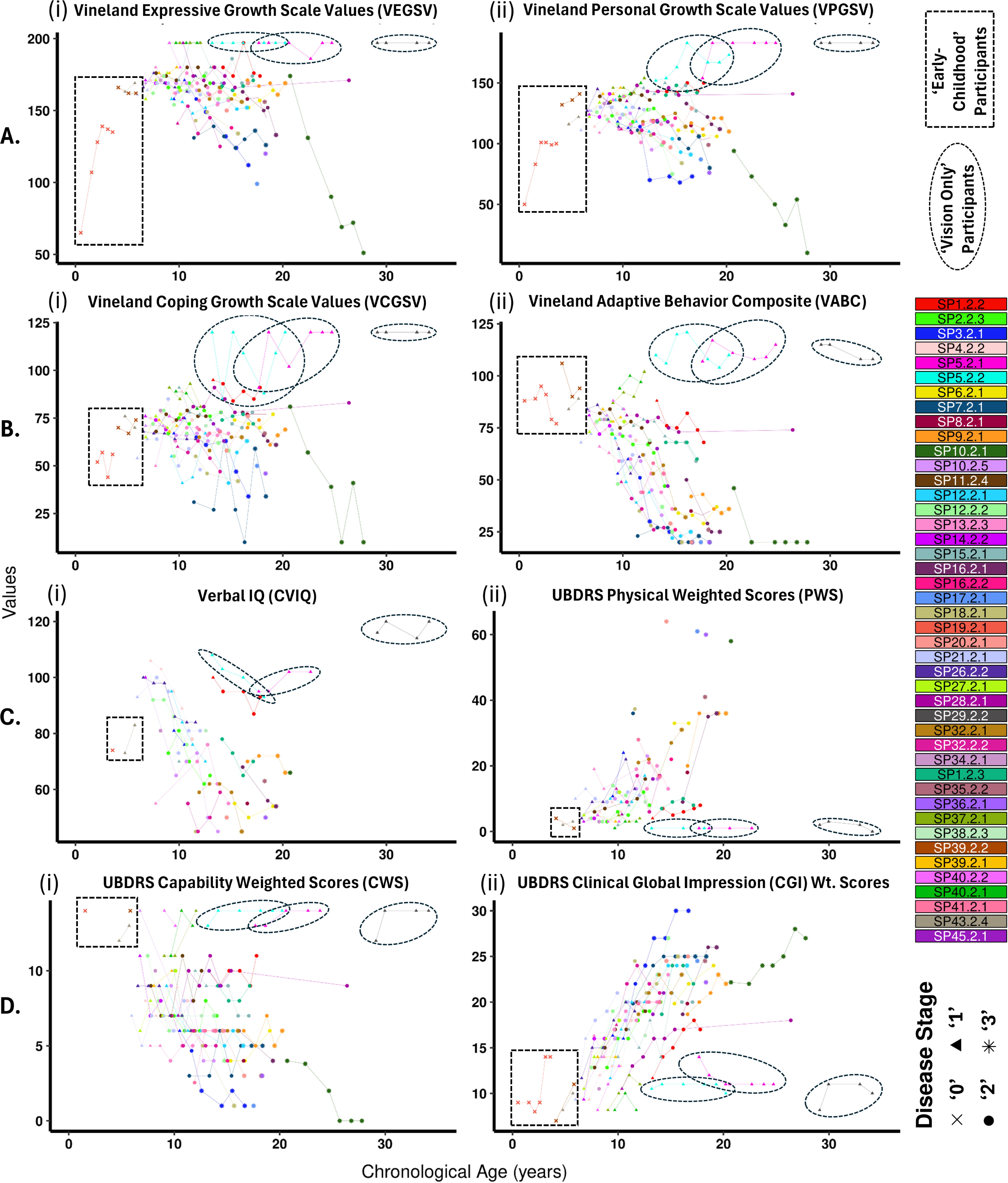
Longitudinal natural history trends in CLN3 individuals for clinical measurements. The generic descriptions in Figure 1 hold for this figure and therefore, not repeated. In general, the Vineland Growth Scale Values (GSV) (**A**(i), **A**(ii) and **B**(i)) tend to show a stronger decline in older participants, albeit with relatively high variability in measurements within participants. The Vineland Adaptive Behavior Composite (VABC) (**B**(ii)) show comparatively less measurement variability within participants and decrease across age more consistently in the syndromic CLN3 individuals. However, VABC tends to show a flooring tendency at later stages (typically, Stage 3) of the CLN3 disease (∼ 18 years and beyond). The VIQ (**C**(i)) measurements show high sensitivity (strong negative trends) up to the age of ∼15 years, and relatively similar flooring effect and measurement variability within participants as VABC (**B**(ii)). The UBDRS Physical scores (**C**(ii)) tend to show stronger increase and higher values in the more severe CLN3 stage (Stage 3), while the UBDRS Capability scores (**D**(i)) show an overall decline with highly variable measurements within individuals. Among all clinical metrics– the UBDRS CGI (**D**(ii)) weighted scores show comparatively more consistent increase in values across age, with the least measurement variability within participants. *Keys:* VIQ = verbal intelligence quotient, UBDRS = Unified Batten Disease Rating Scale, CGI = clinical global impression, Wt. = Weighted.

**Figure 3.**
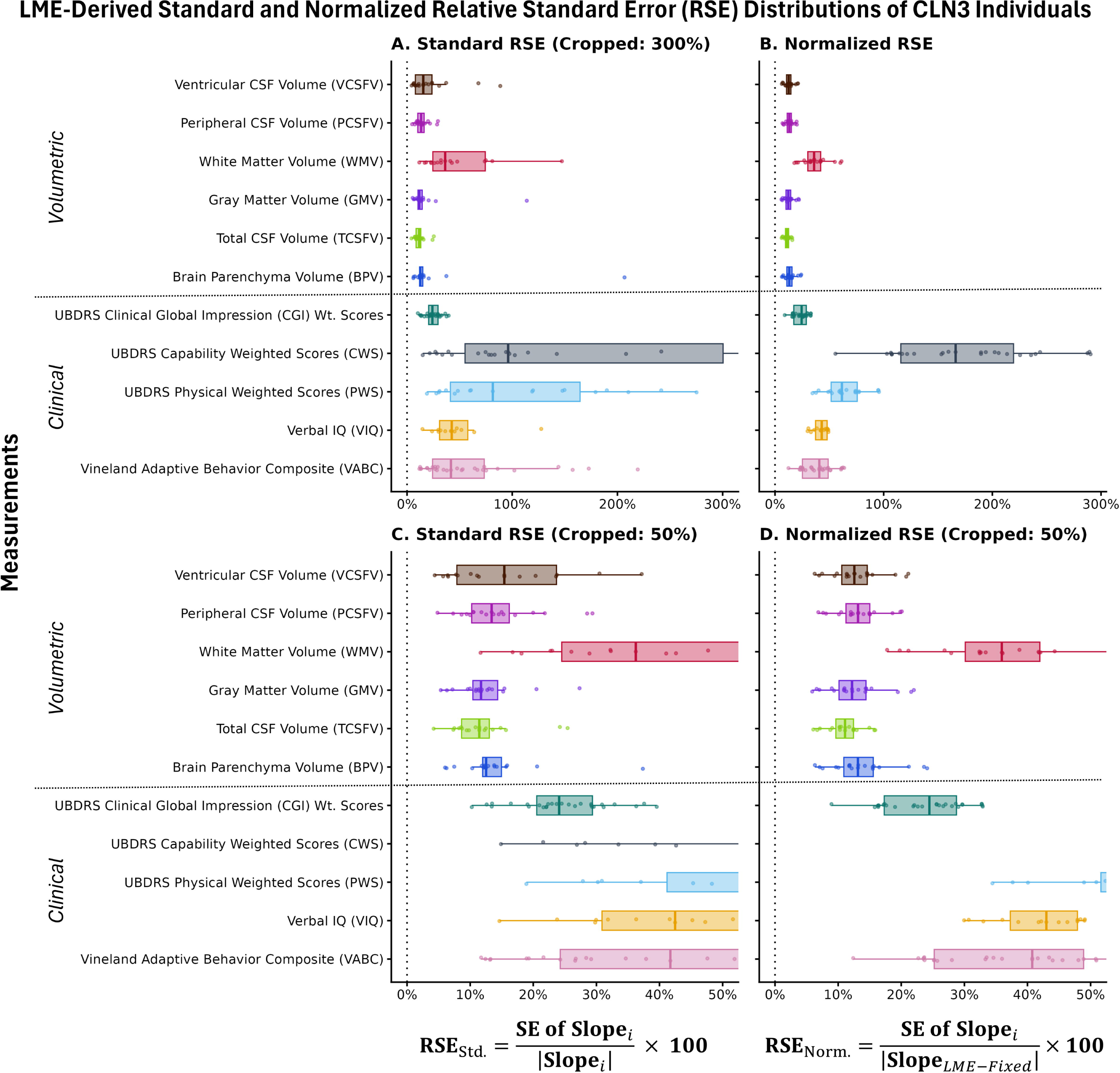
Distributions of standard and normalized relative standard errors (RSEs) across measurements from syndromic CLN3 participants. The y axis represents all measurements, and the dotted horizontal line helps separating the set of volumetric and clinical measurements in all panels. The x axes on the left-side (A, C) represent the standard RSE values, whereas the x axes on the right-side (B, D) represent the normalized RSEs. The x axes titles at the bottom show the mathematical formula used to obtain the values in the left and right panels, respectively. Panel **A** displays boxplots for the standard RSEs of individuals evaluated from the LME model parameters. For an individual *i,* the RSE informs about the level of uncertainty in the slope with respect to its magnitude. This means lower percentages (%) indicate higher precision in the slope of a measurement. The dotted vertical line at x = 0 is provided to assess which measurements tend to be close to the 0% RSE boundary. The plot was cropped at the x = 300% boundary to avoid extreme values distorting the visual. The measurements with tighter (low variability) distributions close to the ‘x = 0’ line can be better appreciated in panel **C**, where the x axis was cropped to show values in the range: 0% – 50% (essentially a ‘zoomed in’ version of panel A). Note that with the exception of the WMV (red), the median RSE of all volumetric measurements tend to be between 10% – 15%, with TCSFV (lime-green) and GMV (violet) showing similar RSEs in the set (∼12%) and BPV very closely following it (∼12.5%). This is significant because it shows that at the individual level, the uncertainty in the MRI-derived atrophy rate is roughly around 12% of the magnitude of the slope, indicating high slope precision. Among the clinical metrics, the UBDRS CGI (dark-teal) has a comparatively lower median RSE (∼24.1%). A critical point to note for the ‘standard RSE’ is that if the magnitude of the slope is 0, or a participant is very stable across their natural history, the standard RSE will tend to go towards infinity (hence the 300% crop-limit in panel A). To avoid such extreme values skewing the distributions, we also showcase a normalized RSE, where the standard error of each random (individual) slope is normalized by the overall average longitudinal slope (LME-derived fixed-effect: *β***_22_**) in the cohort. Panel **B** shows the normalized RSE values across their full range, avoiding extreme RSE values influenced by stable individual slopes and reducing RSE variability between participants (average slope vs. actual individual slope). Panel **D** is a ‘zoomed in’ version of panel B, for a better visual assessment of measurements with low normalized RSEs. *Note:* The *Vineland Growth Scale Values (GSVs –* including *VEGSV, VPGSV,* and *VCGSV)* are not presented here because a stable slope (∼ 0 units/year) is clinically meaningful for these measurements. Because RSE quantification is scaled by the magnitude of the slope, an unchanging slope would lead to a misinterpretation of precision due to highly extreme/infinite RSE values. Consequently, GSVs are excluded here as the RSE would be an invalid measure for these measurements*. Keys:* SE = Standard Error, RSE_Std._ = Standard RSE, RSE_Norm._ = Normalized RSE

### Clinical Measurements

Clinical outcomes were measured via direct assessment, semi-structured caregiver interview, and clinical judgment, in the same week (or rarely within 3 months) as the participant’s MRI scan.

#### Cognitive ability

Due to visual impairments, verbal IQ (VIQ) was the measure of cognitive ability, using the Wechsler system (WPPSI, WISC, or WAIS), yielding age-normed standard scores (population mean=100, SD=15). The test manuals indicate moderately strong correlations between scores on adjacent tests (i.e., WPPSI - WISC and WISC - WAIS). Longitudinal missingness on VIQ may be informative, since cognitive impairment itself may explain why testing was not completed, making the missing-at-random assumption less tenable here than for other performance-based measures.

#### Adaptive behavior

The Vineland Adaptive Behavior Scales (VABS), 3rd Edition is a clinician administered semi-structured caregiver interview yielding an Adaptive Behavior Composite (ABC; mean=100, SD=15) standard score that reflects the individual’s ability to function at an age-appropriate level in the domains of Communication, Daily Living Skills, and Socialization. While Vineland growth scale values (GSVs) are considered more appropriate than standard scores for assessing change,^18^ as an indicator of absolute ability this metric is expected to increase over time at variable rates across development, and so both stagnation and decline may be indications of disease progression. This is incompatible with our statistical methods (see below and eSAP) for evaluating precision and responsiveness, which assume that only change (in the direction of disease worsening) indicates disease progression, and so the ABC remained the focus of this analysis.

CLN3 disease state was assessed using the Unified Batten Disease Rating Scale (UBDRS), which includes four subdomains [Physical, Seizures, Behavior, Capability (with actual vision)] and a Clinical Global Impression (CGI) score. Given the focus here on global brain volumetrics, the primary clinical outcome used was the total weighted CGI score (range: 7 [no signs or symptoms] to 35 [most severe]); Physical and Capability subscores were also evaluated. Disease stage at each visit was assigned per Masten et al.,^19^: ‘0’=pre-symptomatic; ‘1’=vision loss; ‘2’=vision loss+seizures; ‘3’=vision loss+seizures+inability to walk independently.

### Statistical Analysis Plan (SAP)

To address the two fundamental questions posed in the Introduction, each volumetric measurement and COA was evaluated using two complementary metrics derived from their individual-level longitudinal trajectory: the relative standard error (RSE), which quantifies the precision of an individual’s estimated slope relative to its magnitude, and the minimum inter-visit interval required to detect a statistically significant change in that measurement for at least 75% of the cohort (T_min,75%_). Together, these metrics characterize whether a measurement changes in the direction of disease progression on average, and also the precision and responsiveness of the measure. Importantly, the *T_min_* can help guide the minimum follow-up interval future studies should adopt when using a given measurement.

A linear mixed-effects (LME) model framework was used to derive the RSE and T_min,75%_. Briefly, fixed-effects for baseline age and duration in study and a random subject-level intercept and slope of duration were used to predict the outcome. Both RSE and *T_min,75%_* were derived from the random effects of these models. RSE was calculated as the ratio of the standard error (SE) of the slope to the absolute value of the slope. *T_min_* was calculated as a 95% confidence interval around the slope, based on within-participant residual error. The empirical cumulative distribution of *T_min_* across participants was used to identify *T_min,75%_.* The statistical methods are described in detail under Sections 2.1 – 2.3 of the eStatistical Analysis Plan (eSAP) in the eAppendix. A critical point to note – as mentioned earlier under Clinical Measurements, the Vineland GSVs are not compatible with the RSE and *T_min_* framework and therefore, RSE and *T_min_* parameters for these were systematically excluded from related plots and tables.

All statistical analyses were performed in R (version 4.6.1).^20^ As described above, all LME models were fit exclusively within the 38 syndromic CLN3 participants; excluding the ‘early-childhood’ and vision-only participants. Because reliable slope and RSE estimation require a bare-minimum number of repeated observations; participants with fewer than three visits for a given measurement were excluded from that measurement’s LME model. Since there are less MRI than clinical assessments, this restriction produced different effective sample sizes across metrics, all drawn from the same 38-participant cohort. Effective sample sizes, *N* (*≥* 3 visits) for each measurement are reported in Table 2.

**Table 2.** Average Longitudinal Slope (*β*_22_) and its Precision (RSE) for all Measurements.

| Measurement | N ($\geq 3$ Visits) | Overall Average Longitudinal Slope, $\beta_2$ [Bootstrapped 95% CI] | Median [Q1, Q3] Relative Standard Error (RSE) % |
| --- | --- | --- | --- |
| Brain Parenchyma Volume (BPV) | 19 | -1.32 [-1.65, -0.99] | 12.53 [11.98, 14.95] |
| Total CSF Volume (TCSFV) | 19 | 5.67 [4.83, 6.56] | 11.44 [8.64, 13.03] |
| Gray Matter Volume (GMV) | 19 | -1.71 [-2.06, -1.36] | 11.72 [10.43, 14.35] |
| White Matter Volume (WMV) | 19 | -0.66 [-0.98, -0.33] | 36.24 [24.51, 74.38] |
| Peripheral CSF Volume (PCSFV) | 19 | 4.68 [3.92, 5.45] | 13.40 [10.24, 16.20] |
| Ventricular CSF Volume (VCSFV) | 19 | 23.24 [15.75, 30.15] | 15.41 [7.89, 23.67] |
| Vineland Expressive Growth Scale Values (VEGSV) | 29 | -2.22 [-3.48, -0.88]<sup>+</sup> | — |
| Vineland Personal Growth Scale Values (VPGSV) | 29 | -2.08 [-3.49, -0.61]<sup>+</sup> | — |
| Vineland Coping Growth Scale Values (VCGSV) | 29 | -1.36 [-2.27, -0.37]<sup>+</sup> | — |
| Vineland Adaptive Behavior Composite (VABC) | 29 | -3.23 [-4.29, -2.14] | 41.72 [24.28, 73.36] |
| Verbal IQ (VIQ) | 15 | -3.19 [-4.75, -1.61] | 42.47 [30.89, 57.74] |
| UBDRS Physical Weighted Scores<br>(PWS) | 19 | 1.50 [0.51, 2.51] | 81.54 [41.19, 164.46] |
| UBDRS Capability Weighted<br>Scores (CWS) | 27 | -0.15 [-0.39, 0.09] | 96.09 [55.11, 300.42] |
| UBDRS Clinical Global Impression<br>(CGI) Weighted Scores | 28 | 1.26 [1.03, 1.49] | 24.10 [20.55, 29.38] |

‡: For the Vineland Growth Scale Values (GSVs) – since a significant interaction between the baseline age and longitudinal age was observed – the average longitudinal slope was computed after accounting for the interaction term (see eq.[5] in eSAP). Also, because RSE is not a valid metric for the GSVs – they were left empty. For all other measurements – the longitudinal age effect was separately characterized from the between-participants baseline effect. Therefore, they were tabulated directly from the $\beta_2$ estimates (see eq.[4] in eSAP) from their LME models. The 95% confidence intervals (CIs) for all measurements were obtained by bootstrap resampling for better accuracy and robustness. For RSEs – the median and the inter-quartile values [Q1, Q3] are shown, because several measurements have highly extreme values at the tails. Keys: **N ( $\geq 3$ Visits)**: the effective sample size that includes only participants with at least 3 or more visits out of the 38 syndromic CLN3 participants.

### Artificial Intelligence Disclosure

The first author wrote the entire first draft of the manuscript with ‘no use of AI’. Co-authors provided feedback and edits. A government approved version of Claude (Sonnet 5; Anthropic; accessed via claude.hhs.gov) was then used to help consolidating the inputs from all authors and improving clarity and readability of the Introduction and Discussion sections. All AI-generated texts were reviewed, and if necessary, further edited by the authors, who take full responsibility for the accuracy and content of the final manuscript.

## RESULTS

### Baseline Characteristics

Baseline characteristics of the 38 CLN3 participants included in formal statistical analyses are summarized in Table 1. Participants had a mean baseline age of 11.5 years (SD 4.3; median 10.2, range 6.0–20.7) at their first MRI scan, with equal representation by sex (19 female, 19 male). Most participants carried the common 966-base-pair deletion, either homozygous (n=17, 45%) or as compound heterozygous (n=16, 42%); the remaining 5 participants (13%) carried other genotypes. At baseline, 16 (42%) were at Stage ‘1’, 17 (45%) at Stage ‘2’, and 5 (13%) at Stage ‘3’.

Baseline volumetric measurements were complete for all 38 participants. Baseline clinical measurements showed varying degrees of missingness: Vineland GSVs and Vineland ABC (VABC)were each missing for 1 participant, UBDRS Capability score was missing for 1 participant, and UBDRS Physical and CGI were complete for all participants. VIQ had the highest missingness at baseline (12 missing – 11 due to English not being the particiapants’ primary language).

### Longitudinal Volumetric and Clinical Measurements

Unmodeled longitudinal trajectories of all volumetric measurements showed consistent patterns of progressive brain atrophy across the syndromic CLN3 cohort (Figure 1). BPV and GMV declined steadily with age and showed relatively low measurement variability, while WMV declined more modestly and with greater variability. Consistent with the Monro-Kellie doctrine, the total CSF volume (TCSFV) increased with age, compensating for parenchymal tissue loss. The constituent peripheral and ventricular CSF volumes followed a similar pattern but with relatively higher variability. Early-childhood and vision-only participants were visually distinct from the syndromic CLN3 trajectory in all panels, supporting their exclusion from formal modeling.

Raw clinical measurements showed more heterogeneous trends (Figure 2) than volumetric measurements (Figure 1). Vineland GSVs (Expressive, Personal, Coping) declined more steeply in older participants but with high within-participant variability; the VABC declined more consistently, though with an apparent flooring effect beyond approximately 18 years of age (mostly, Stage ‘3’). VIQ showed strong negative trends up to approximately age 15, with a similar flooring pattern thereafter. Among UBDRS subscales, the Physical score increased and the Capability score decreased with age, both with substantial within-participant variability; the CGI showed the most consistent increase with age and the lowest within-participant variability among all clinical measures.

Two participants included in the syndromic CLN3 cohort showed individual trajectories worth noting (eAppendix: eFigures 4, 5). SP13.2.3 had a congenital arachnoid cyst that was misclassified by SynthSeg as peripheral CSF, inflating TIV; despite this, the participant’s brain parenchyma showed clear, progressive atrophy across visits, consistent with the overall cohort trend. SP40.2.1 showed a relatively stable BPV trajectory across three visits with minimal evidence of atrophy, more closely resembling a typically developing brain. This participant’s CLN3 genotype has not been previously reported in the literature; therefore, while the participant’s vision loss follows the typical time course other neurological presentations may not.

**Figure 4.**
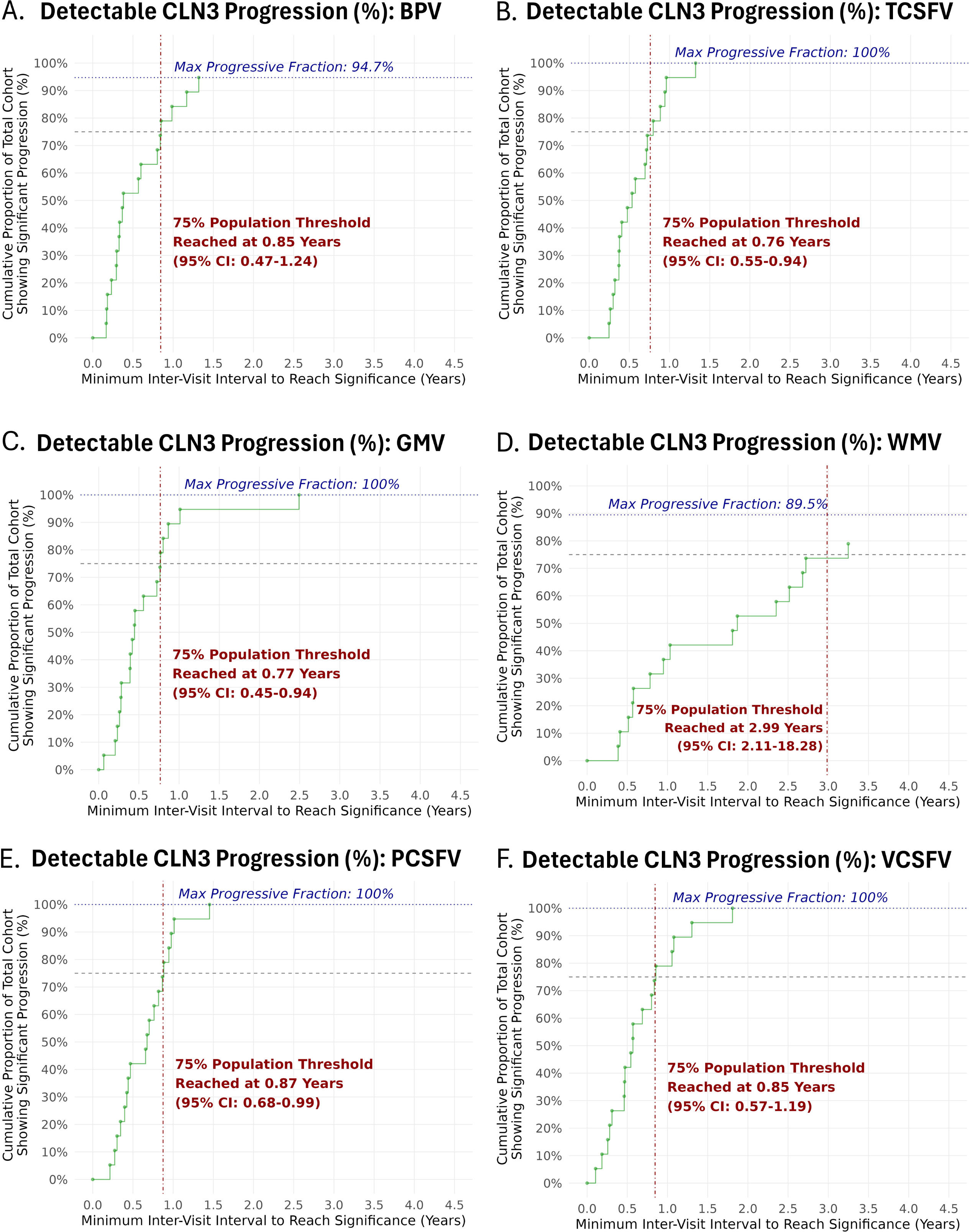
Distribution of minimum inter-visit interval (*T_min_*) for volumetric measurements to document significant progression in the CLN3 cohort. The figure shows the empirical cumulative distribution function (ECDF) of each volumetric measurement against the minimum inter-visit interval times (*T_min_*) required for CLN3 individuals to undergo a significant volume change towards neurodegeneration. The x axis is cropped at 4.5 years for all measurements because an interval longer than 4 years for 75% of the population to reach significance (*T_min,75%_*) – is practically very long to be clinically useful. The y axis represents the percentage of the entire cohort that reaches significance for a certain *T_min_*. From an experimental design and clinical trial perspective, this can be very informative. For example, the ECDF of the brain parenchyma volume (A: BPV) shows that 75% of the cohort reaches significance for a minimum inter-visit interval of 0.85 years. This suggests that future observational study or clinical trials should consider tracking CLN3 participants for at least 0.85 years to guarantee that approximately 75% of the population demonstrate neurodegeneration in CLN3 disease. The 75% threshold is arbitrary, and it depends on the investigator’s prerogative to decide what proportion they consider to be valid to document progression. Based on the current study design – repeating MRI visits annually (i.e., for a *T_min_* of 1 year) may guarantee that CLN3 progression can be reliably documented in at least 85% of the population (obtained from the BPV distribution in A). Note, it is entirely possible that not 100% of the population will reach significance, if, for example, certain individuals have stable or positive slopes not aligning with disease progression. For the BPV in panel A, the maximum progressive fraction that can be reached is 94.7% because a CLN3 individual has a slight positive trend. In the case, where some individuals have slopes with very small magnitude, the *T_min_* will be naturally longer – the change in volume over a shorter interval may not be strong enough to overcome the noise boundary. This can be observed for the WMV in panel **D**, where the *T_min_* is ∼ 3 years for 75% of the cohort to reach significance. For measurements with similar error levels, a stronger slope will require shorter intervals. For example, TCSFV (panel **B**) and GMV (panel **C**) show very similar *T_min,75%_* (0.76 and 0.77 years, respectively) understandably because the increase in TCSFV majorly offsets the loss in GMV (the most atrophic tissue). Panels **E** and **F** show the distributions for the peripheral and ventricular CSF volumes, respectively, which comprise the TCSFV. Note the slightly higher *T_min,75%_* for these two CSF constituents (0.87 and 0.85, respectively) which may reflect the higher measurement error arising from measuring volumes in smaller compartments. *Keys:* BPV = Brain Parenchyma Volume, **TCSFV** = Total CSF Volume, **GMV** = Gray Matter Volume, **WMV** = White Matter Volume, **PCSFV** = Peripheral CSF Volume, **VCSFV** = Ventricular CSF Volume, Minimum Inter-Visit Interval (*T_min, i_*) for the *Slope* of a participant (*i*) to reach significance was computed by: 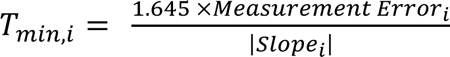

### LME-Modeled Longitudinal Trends

Individual-level LME model fits confirmed the longitudinal patterns observed in the raw data described above. All volumetric measurements showed statistically significant longitudinal slopes (eFigure 6 and Table 2), notably: BPV (−1.32%/year, 95% CI: −1.65 to −0.99), GMV (−1.71%/year, 95% CI: −2.06 to −1.36), and WMV (−0.66%/year, 95% CI: −0.98 to −0.33) all declined significantly with age, while TCSFV (5.67%/year, 95% CI: 4.83-6.56) increased significantly, consistent with compensatory CSF expansion.

Among clinical measures, all showed statistically significant longitudinal slopes (eFigure 7 and Table 2) except UBDRS Capability, whose 95% CI crossed zero (−0.15/year, 95% CI −0.39 to 0.09) despite the slope moving in a direction consistent with expected disease progression (declining capability). UBDRS CGI showed the most consistent clinical effect (1.26 units/year, 95% CI 1.03–1.49).

### Precision of Individual-Level Slopes (RSE)

Relative standard error (RSE) distributions (Figure 3) showed higher precisions in the volumetric measurements than in the COAs at the individual level. With the exception of WMV, all volumetric measures had median RSE between 10%–15%, with BPV, TCSFV and GMV showing comparable RSEs of roughly around 12% (Table 2). Among clinical measures, UBDRS CGI showed the lowest median RSE (∼24.1%), though showing substantially less overall precision than the BPV (∼12.5%).

### Minimum Inter-Visit Interval (*T_min_*)

Empirical cumulative distribution functions of *T_min_* (Figures 4, 5) identified the minimum follow-up interval needed to detect significant longitudinal change in a given proportion of the cohort. Among volumetric measures, TCSFV and GMV required similar *T_min,75%_* of ∼0.80 years, followed closely by BPV (0.85 years, 95% CI 0.47–1.24). Although GMV’s *T_min,75%_* was marginally shorter, BPV was prioritized as the primary volumetric candidate throughout this study given its reduced susceptibility to segmentation error in pathological brains relative to isolated GM or WM measurements (see eMethods); the two measures otherwise performed comparably on both RSE and *T_min_*. WMV required a substantially longer interval (∼3 years) to reach the same threshold, consistent with its smaller effect size and higher variability.

UBDRS CGI (Figure 5A) showed the shortest *T_min_* among all clinical measurements (*T_min,75%_* = 2.56 years; *T_min,50%_* = 1.5 years for 50%). VIQ and VABC (Figure 5 B, C) required longer intervals to reach 75% significance (3.5 and 4.0 years, respectively) but could each document progression in 50% of the cohort within 2 years. UBDRS Physical showed a *T_min_* profile similar to VABC. UBDRS CWS did not reach the 75% threshold – likely due to high inter-visit measurement variability.

**Figure 5.**
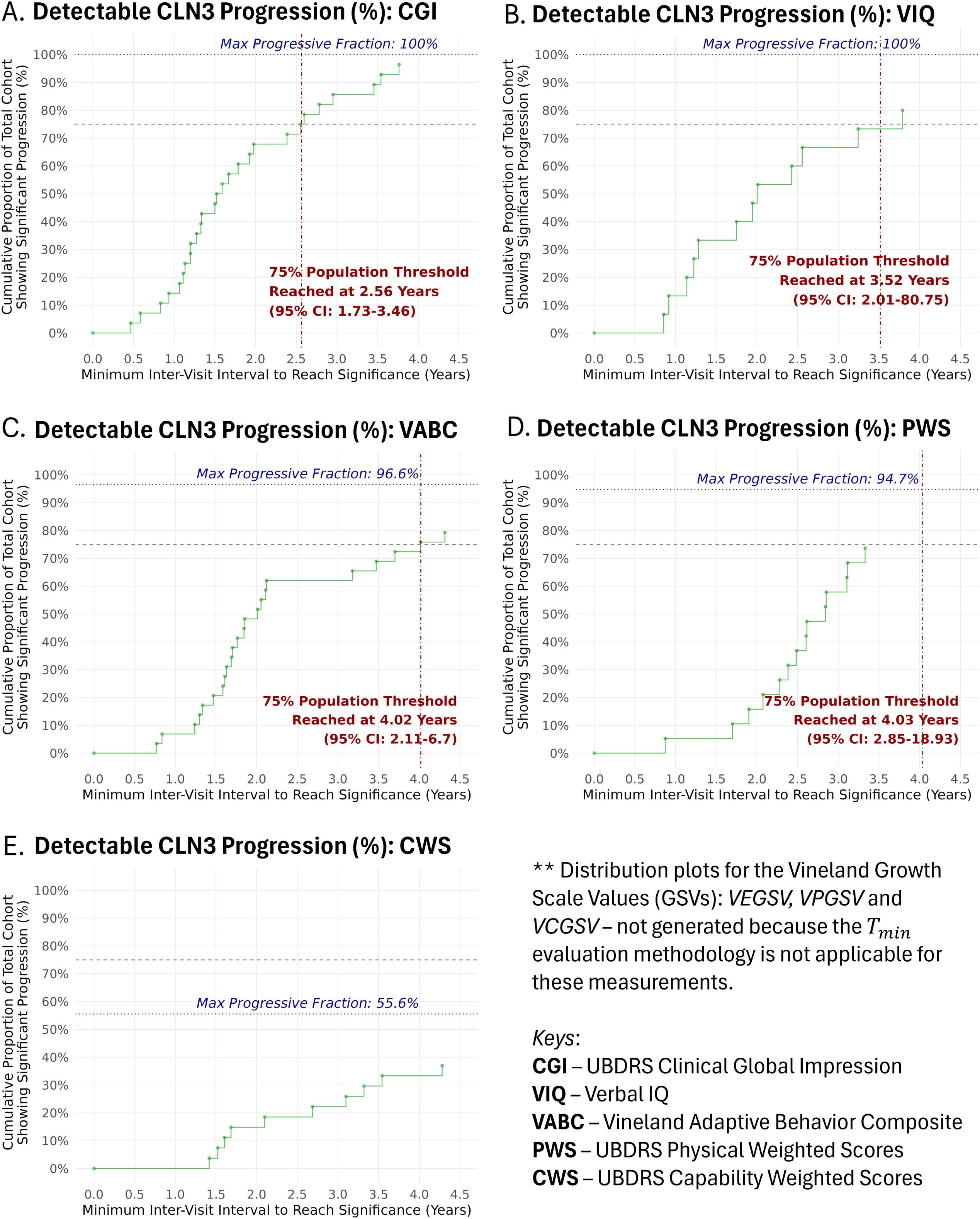
Minimum inter-visit interval for clinical measurements to document significant progression in the CLN3 cohort. The generic descriptions in Figure 4 apply here and therefore are not repeated. For the purposes to highlight the most optimal candidates, the measurements are shown in order of lowest to highest *T_min,75%_,* to reach the 75% cohort significance. The UBDRS CGI (**A**) showed the smallest *T_min,75%_* (2.56 years) among all clinical measurements. This is still a relatively longer interval for tracking CLN3 progression; however, the distribution suggests that a *T_min,50%_* of ∼1.5 years may detect progression in 50% of the population. Similarly, for VIQ and VABC (Panels **B** and **C**, respectively), although the *T_min,75%_* is relatively longer (3.52 and 4.02 years), they both can be used to document significant progression in 50% of the population with a *T_min,50%_* of < 2 years. Note, because the x-axis was cropped at the 4.50 years boundary, participants with much longer *T_min_* are not visible. Theoretically, given sufficient longitudinal tracking, both UBDRS CGI and VIQ can capture progression in 100% of the cohort, though this is less practical in standard clinical trial settings.. The distribution for UBDRS PWS is shown in panel **D,** which tends to have a similar *T_min_* as VABC for the 75% threshold. For the UBDRS CWS measurements (**E**), the *T_min_* values were the longest in the set and never reached the 75% threshold – likely due to high inter-visit variability in this measurement. *Note:* The *Vineland Growth Scale Value (GSV)* distributions are excluded here. This is mainly because the *T_min_* methodology relies on a change of measurement in the direction of disease progression. For the Vineland GSVs – even a ‘no change’ (highly stable or slope = 0) may indicate progression because healthy, normative growth values are expected to increase with age. Therefore, the *T_min_* computation is not valid for these measurements.

## DISCUSSION

To support clinical trial readiness, we evaluated the precision and responsiveness of MRI-derived brain volumetric measures and several COAs in CLN3 disease in this study. To do so, we also resolved several methodological questions about whether and how MRI-derived measures reflect true brain atrophy in this population, rather than be artifacts of segmentation or normalization.

A quantitative biomarker of atrophy is only as reliable as the data and processing pipeline used to derive it. Both data quality and segmentation software choice were essential considerations here, given that CLN3 produces progressive, pathology-driven changes to brain structure and tissue contrast that differ from the typically developing and healthy adult brains on which most segmentation tools are validated. Total intracranial volume (TIV) is generally assumed to stabilize after approximately age three,^21^ an assumption that underlies the conventional practice of normalizing volumetric measures by TIV. We confirmed this assumption held using several segmentation tools in MRI-data from typically developing children’s datasets^22, 23^ (eMethods: Section 1.3) within our age range of interest. Paradoxically, however, TIV as estimated by the same tools showed a clear decline with age within our CLN3 cohort (eFigure 3A(ii)); the magnitude of this artifactual decline varied by algorithm, but no tool we tested was immune. This indicates that the effect reflects a segmentation confound specifically induced by CLN3 pathology, rather than a general limitation of pediatric TIV estimation. Even contemporary, AI-based segmentation tools such as SynthSeg – designed to be robust to variation in acquisition contrast and resolution – may not fully generalize to a progressively atrophying, pathological brain. This finding has a broader methodological implication: segmentation tools validated in adults or typically developing children may require independent validation before being applied to disease populations with atypical structural or contrast features, since pathology-driven confounds are not guaranteed to be caught by tools designed for anatomical robustness alone. Because no available tool was immune to this bias, we developed the LME-based TIV adjustment described in eMethods (Section 1.2) specifically to correct for it; left uncorrected, this bias would have led to a systematic underestimation of atrophy (eFigure 3C(i) vs. C(ii)).

With a corrected, individual-specific baseline TIV established, we next considered which volumetric compartment should serve as the primary biomarker of atrophy. GMV, BPV and TCSFV in particular – all showed strong and broadly comparable precision and responsiveness (median RSE ∼12%, *T_min,75%_* < 1 year; Table 2 and Figures 3, 4), consistent with the fact that parenchymal loss and CSF expansion are, by construction, complementary reflections of the same atrophic process and BPV necessarily carry equivalent information about the parenchyma-CSF boundary.

Despite this broad statistical equivalence, several considerations favor BPV as the preferred general-purpose metric. First, GMV, WMV, PCSFV, and VCSFV each require a finer level of tissue classification than BPV: distinguishing gray from white matter, or peripheral from ventricular CSF, requires resolving boundaries that are more susceptible to pathology-driven contrast changes^24, 25^ and to the specific assumptions of the segmentation algorithm used, whereas BPV requires only the single, higher-contrast parenchyma-CSF boundary. Second, and specific to the CSF-based measures, both PCSFV and VCSFV are comparatively small-volume compartments that show substantial inter-individual variability even in typically developing children, making it inherently more difficult to establish a stable normative reference value against which to judge atrophy; BPV, drawing on the much larger and comparatively more consistent parenchymal compartment, is less exposed to this problem. Taken together, we consider BPV likely to be the preferable general-purpose metric for multicenter or multi-platform studies, given its reduced sensitivity to GM/WM segmentation method, its comparatively lower dependence on the specific acquisition sequence used, and its lower inter-individual variability in normative values relative to the CSF-based measures – notwithstanding GMV’s marginally better statistical performance within this single-site, single-pipeline dataset (median RSE 11.7% vs. 12.5%; *T_min,75%_* of 0.77 vs. 0.85 years). WMV remained the least robust volumetric measure on every axis (*T_min,75%_* ≈ 3 years; Figure 4D), consistent with both a smaller true effect size and this same finer-grained segmentation vulnerability.

Two individual trajectories further illustrate both the robustness and the limits of this framework. One participant with a congenital arachnoid cyst (SP13.2.3) showed SynthSeg misclassification of cyst fluid as peripheral CSF, inflating TIV, yet still displayed clear, progressive parenchymal atrophy consistent with the overall cohort trend – suggesting BPV remained informative despite a segmentation artifact affecting a different compartment.

Conversely, one participant with a previously unreported CLN3 genotype (SP40.2.1) showed a stable BPV trajectory across three visits that more closely resembled typical development; this participant represents the sole case preventing BPV’s maximum progressive fraction from reaching 100% (94.7%; Figure 4A) and raises the possibility of genotype-related heterogeneity in the rate, if not the eventual occurrence, of neurodegeneration in CLN3.

Our normalization approach also established a meaningful normative reference for “no atrophy” (BPV = 100%) using healthy pediatric reference data.^22, 23^ Notably, the youngest syndromic CLN3 participants (∼5–6 years of age) showed a parenchymal fraction similar to that expected in typically developing children of the same age (eFigure 3C(ii)) – reassuring evidence that substantial atrophy has not yet occurred at this stage, and one that reinforces the potential value of early diagnosis should an effective neuroprotective treatment become available. This is consistent with the older, syndromic CLN3 cohort analyzed here overall: mean BPV at a mean baseline age of 11.5 years was 94.9% of the healthy expected volume (Table 1), indicating that meaningful parenchymal loss accumulates progressively over childhood and adolescence.

On average, syndromic CLN3 participants lost approximately 1.3% BPV, 1.7% GMV, and 0.7% WMV per year. This is directionally consistent with, but numerically lower than, GMV and WMV atrophy rates reported in a prior CLN3 natural history study (4.6% and 1.0% per year, respectively, measured from supratentorial GM/WM only – excluding cerebellum and brainstem).^16^ Aside from the difference in tissue exclusion (cerebellum and brainstem included in this study) – this discrepancy may reflect differences in MRI acquisition, processing, and analysis – including image contrast, segmentation methodology, normalization approach, and time-dependence modeling – as well as differences in cohort composition, underscoring the broader point that methodological choices materially affect atrophy estimates and should be reported and compared explicitly across studies.

Having established BPV as a reliable, methodologically robust measure of atrophy, we turned our attention to COAs commonly used for CLN3. UBDRS CGI, VABC, and VIQ are behaviorally anchored measures: they depend on a child’s engagement, caregiver report, clinical interpretation, and – particularly at more advanced disease stages – normative floor effects that compress the dynamic range available to detect further decline. We observed this directly: for VABC in particular – trajectories flattened beyond approximately 15–18 years of age, largely coincident with Stage 3 disease (Figure 2B(ii)). Conversely, MRI volumetrics showed no equivalent floor within the age and disease-severity range studied here, and they exhibit far greater reliability and responsiveness. We note that the valid interpretation of RSE and *T_min_* rests on disease progression being expressed as a slope that differs from zero. For certain types of scores, such as the Vineland GSVs, a *lack* of change may indicate failure to achieve developmental expectations and therefore disease progression. The clinical trial estimand for this type of score is the difference in two slopes (within or between-subject), rather than the slope itself. Thus, the interpretation of RSE and *T_min_* for this type of score is not valid, and we did not include them here. We also recognize that psychometric problems, such as normative floor effects in standard scores like VIQ, can impact the validity of the RSE and *T_min_* by failing to reflect variability below an arbitrary score.

From a trial design perspective, the *T_min_* in these COAs was comparatively long. The UBDRS CGI reached the 75% threshold sooner than all other COAs, though both VIQ and VABC could document progression in 50% of the cohort within 2 years (Figure 5B-5C). This limited responsiveness underscores that clinic-based instruments may require substantially longer follow-up, larger samples, or both, to reliably power an early interventional trial in this rare population. Consequently, trial design must carefully balance the trade-off between statistical reliability and clinical validity when selecting a COA. For example, while VIQ and VABC demonstrate lower reliability (longer *T_min_* intervals and higher RSEs) than the UBDRS CGI, their capacity to assess domain-specific cognitive and adaptive functions in CLN3 may outweigh these statistical disadvantages. However, for VABC, the clinical validity is particularly nuanced^18^ because observed declines can reflect either decelerated developmental gains or absolute functional deterioration – two trajectories with vastly different clinical interpretations. These operational factors must be factored into COA selection for prospective clinical trials.

These results carry direct implications for trial design in CLN3 disease. A treatment expected to slow the natural trajectory of BPV could plausibly demonstrate a detectable signal in well under a year per participant using the framework applied here – indeed, annual MRI, the interval used in this study, was sufficient to capture significant atrophy in approximately 85% of the cohort (Figure 4A) – whereas relying on UBDRS CGI alone would require a considerably longer observation period for comparable confidence, and other COAs longer still. Because individual BPV trajectories tracked the population-level trend closely, a newly diagnosed CLN3 participant can also reasonably be expected to follow a similar atrophy trajectory if the genotype is consistent with a syndromic presentation, which may aid individualized clinical assessment and counseling in addition to its trial-design implications. While additional work is needed to establish the predictive relationship between volumetric measures and meaningful aspects of health, the results of this study supporting the reliability and responsiveness of MRI-derived BPV position it as a strong co-primary candidate endpoint in future interventional studies along with clinical measures – particularly UBDRS CGI – that capture clinically meaningful, rather than purely biological, benefit.

### Limitations

The precision of the slope and RSE parameters estimated here depends on the number of longitudinal observations available per participant. Approximately one-quarter of participants included in the volumetric analyses had only three MRI visits – the bare minimum required for the individual-level LME estimation used throughout this study – and estimates for these participants are inherently less stable than for participants with longer follow-up. Effective sample sizes also varied across measurements (Table 2) due to differing visit-completion patterns; this likely most affected measures with higher percentage of missing values such as VIQ, which also carried informative missingness from participants too impaired to complete testing. MRI and clinical assessments were not always obtained on the same day, introducing a small amount of temporal misalignment. This was an observational longitudinal study; we cannot yet confirm that treatment-related slowing of BPV decline corresponds to clinically meaningful benefit, only that the measure is well suited to detecting a biological signal if one exists. Early-childhood and vision-only participants were excluded from formal modeling given their qualitatively distinct trajectories (eFigure 3 and Figures 1–2); these small subgroups may nonetheless be informative for future work on presymptomatic or non-syndromic biomarkers, and their exclusion here should not be read as evidence that MRI volumetrics are uninformative in these disease courses.

### Generalizability

The high precision and responsiveness we observed for volumetric measurements in this cohort were achieved using a single, high-resolution 3T T1w acquisition protocol and a single segmentation pipeline (SynthSeg v2.0), in a cohort scanned under sedation to minimize motion artifact. Reliability could be lower in cohorts with more heterogeneous acquisition parameters, shorter or lower-resolution sequences, greater motion artifact, or different field strengths, and could plausibly vary with the segmentation software used, given the pathology-related segmentation confounds discussed above. While BPV’s reduced dependence on fine-grained tissue classification should make it comparatively more robust to these sources of variability than GMV, WMV, or the individual CSF compartments, this has not been directly tested across sites or protocols, and multicenter validation would be needed before generalizing the specific precision and *T_min_* values reported here to other acquisition or processing environments.

## Conclusion

MRI-derived brain parenchyma volume, and by extension its mathematical counterpart total CSF volume, are highly precise and responsive measurements for tracking progressive neurodegeneration in CLN3 disease – detecting reliable, individual-level change within a follow-up interval well suited to trial timelines in a rare disease population. Incorporating these volumetric measures alongside clinical assessment – rather than relying on either in isolation – offers the most efficient path toward adequately powered, feasibly timed clinical trials of emerging CLN3 therapies.

## Supporting information

eAppendix

## Data Availability

De-identified and anonymized individual-level participant data supporting the findings of this study have been compiled into a dedicated spreadsheet named eTable 1 (Excel format). To ensure full compliance with target journal guidelines, this comprehensive minimum dataset will be made openly accessible to qualified researchers within the online supplemental material immediately following official peer-reviewed publication.

To support the independent reproduction and verification of our results in alignment with Good Clinical Practice guidelines, all underlying data have been prepared for open access. De-identified longitudinal individual participant demographics, raw clinical outcome assessments, and MRI-derived normalized brain measurements across all study visits are compiled in eTable 1. This file is designated for public release via the online supplemental material immediately upon peer-reviewed journal publication.

## Acknowledgments

1. This research was supported by the Intramural Research Program of the NIH (Z1A HD009001 to ADD, ZEB000088-10 to CP).
2. Data used in the independent analysis of computing brain parenchyma fraction in typically developing children (N = 121) were obtained from the <u>Adolescent Brain Cognitive Development™ (ABCD) Study</u>, held in the <u>NIH Brain Development Cohorts Data Sharing Platform</u>. This is a multisite, longitudinal study designed to recruit more than 10,000 children aged 9–10 and follow them over 10 years into early adulthood. The ABCD Study® is supported by the **National Institutes of Health** and additional federal partners under award numbers: U01DA041048, U01DA050989, U01DA051016, U01DA041022, U01DA051018, U 01DA051037, U01DA050987, U01DA041174, U01DA041106, U01DA041117, U01 DA041028, U01DA041134, U01DA050988, U01DA051039, U01DA041156, U01D A041025, U01DA041120, U01DA051038, U01DA041148, U01DA041093, U01DA0 41089, U24DA041123, U24DA041147. A full list of supporters is available at Federal Partners – ABCD Study. ABCD Consortium investigators designed and implemented the study and/or provided data but did not necessarily participate in the analysis or writing of this report. This manuscript reflects the views of the authors and may not reflect the opinions or views of the NIH or ABCD Consortium investigators.
3. We value the continued motivation and inspiration provided by study participants and their families and appreciate their dedication to help move research forward. Special thanks to Murdoch Childrens Research Institute, Melbourne, Australia and the Department of Pediatrics, University of Melbourne, Melbourne, Australia for facilitating the data sharing from Neuroimaging of the Children’s Attention Project (NICAP) study. This was the second independent database (N = 38) used to compute the brain parenchyma fraction in healthy children. This study was supported by the National Health and Medical Research Council (NHMRC) of Australia (Project Grant No. 1065895). We appreciate the Murdoch Children’s Research Institute and The Royal Children’s Hospital’s Medical Imaging staff for their assistance and expertise in MRI data collection.

## Grant Support

None

## DISCLOSURE STATEMENTS

**USU:** “The opinions and assertions expressed herein are those of the author(s) and do not reflect the official policy or position of the Uniformed Services University of the Health Sciences or the Department of War.”

**HJF:** “The view(s) expressed herein are those of the author(s) and do not reflect the official policy or position of the Henry M. Jackson Foundation for the Advancement of Military Medicine Inc. (HJF).”

**NIH:** This research was supported by the Intramural Research Program of the National Institutes of Health (NIH). The contributions of the NIH authors were made as part of their official duties as NIH federal employees, are in compliance with agency policy requirements, and are considered Works of the United States Government. The findings and conclusions presented in this paper are those of the authors and do not necessarily reflect the views of the NIH or the U.S. Department of Health and Human Services.

## SUPPLEMENTAL MATERIAL CONTENTS

**eAppendix.** Comprehensive methodological descriptions, especially those involving the brain volumetric data, statistical analysis plan, and ancillary analytical figures (PDF format).

**eTable 1.** Database comprising longitudinal Participant Demographics, Clinical Outcome Assessment measurements and MRI-Derived Normalized Brain Compartmental Volumes across all study visits. This Microsoft Excel spreadsheet contains the row-by-row, de-identified minimum dataset optimized for reproducibility.

- **Sheet 1 (eTable_Legend):** Explicit definitions for all column variables, acronyms, coding schemas and units of measurement.
- **Sheet 2 (eTable):** Individual-level longitudinal participant tracking including demographics, raw clinical outcome assessments and normalized brain compartment volumes derived using MRI.

*Note:* This dataset is designated for public release in accordance with official journal guidelines immediately following peer-reviewed publication approval.

## Notes

### Competing Interest Statement

The authors have declared no competing interest.

### Author Declarations

Recruitment and evaluation were approved by the NIH Institutional Review Board and performed per International Conference on Harmonization Good Clinical Practice guidelines and applicable US Code of Federal Regulations.

