## Supplementary material for "Evaluating Precision and Responsiveness of MRI-derived Brain Volumetric and Clinical Outcome Measures in CLN3 Disease": eAppendix

#### 1.0 eMethods

#### 1.1 SynthSeg Segmentation

We evaluated several contemporary segmentation and brain-extraction tools across healthy and pathological pediatric imaging databases to identify the most appropriate method for this cohort; those comparative results are outside the scope of this manuscript and will be reported separately. SynthSeg was selected for its reproducibility and robustness relative to the other tools tested. 'SynthSeg' uses 3D U-Net, a convolutional neural network architecture suited to segmenting 3D brain images.[1, 2] It was designed to be robust to variation in image contrast, resolution, and artifacts across acquisition protocols. Now distributed as part of FreeSurfer (http://surfer.nmr.mgh.harvard.edu/)[3-6], it can generate volumetric outputs matching the standard FreeSurfer segmentation labels. We used the SynthSeg version 2.0 within FreeSurfer (version 7.4.1) to segment all T1w images independently in their image-reconstructed native space.

eFigure 2 illustrates SynthSeg segmentation output for one participant at their first and fifth visits, showing visible atrophy alongside the expected age-related increase in head size. Total intracranial volume (TIV), by contrast, remained essentially constant between visits, while peripheral and ventricular CSF volume increased– consistent with the Monro-Kellie doctrine, which holds that once cranial growth is complete (typically after age 3) and TIV stabilizes,[7] any loss of parenchyma volume must be offset by a corresponding increase in CSF volume.[8, 9]

**1.2 Computation of Brain Parenchyma Volume**

SynthSeg outputs raw volumes for individual brain regions and compartments, along with TIV, computed as the sum of all segmented volumes within the cranial cavity. Raw brain parenchyma volume (BPV) was computed as the sum of all cortical, subcortical, white matter, brainstem, and cerebellar label volumes. Gray matter (GM), white matter (WM), and CSF volumes were each computed analogously. BPV was chosen as the primary volumetric outcome, rather than GM or WM separately, because it is less susceptible to segmentation errors arising from pathological changes in a diseased brain.[10, 11] The same normalization and analytic pipeline described below was also applied separately to each tissue compartment. As a validation check, we confirmed that the raw BPV was equivalent to the raw TIV minus the raw total CSF volume – the sum of the raw peripheral and ventricular CSF volumes (PCSFV and VCSFV, respectively) at the individual level, using equation [1] below:

$$Raw\ BPV_{i,k} \equiv TIV_{i,k} - TCSFV_{i,k} \equiv TIV_{i,k} - (PCSFV_{i,k} + VCSFV_{i,k}) \quad [1]$$

where, $i = 1, 2, 3 \ldots N$ subjects and $k = 1, 2, 3 \ldots M$ visits.

**1.3 Normalization of BPV**

When raw BPV and TIV were plotted against age, nearly all participants showed consistent negative BPV slopes– evidence of atrophy even before normalization (eFigure 3A). TIV also declined with age, though less steeply than BPV; except in the 'pre-symptomatic' and vision-only participants who showed positive and stable TIV trajectories, respectively.

Raw BPVs are conventionally normalized by each participant's TIV (ratio method), though model-based residual normalization is often preferred.[12, 13] The ratio method works, when TIVs are longitudinally stable across age; because TIV instead declined with age in this cohort–

likely reflecting disease-related interference with segmentation, rather than true anatomical change– a simple ratio would underestimate atrophy (less steeper slopes in eFigure 3C(i)).

TIV is a useful proxy for brain volume at a neurologically healthy baseline, before neurodegeneration begins. To correct for the disease-related decline in measured TIV, we fit a linear mixed-effects (LME) model with each individual’s longitudinal age centered by their baseline age, so the model intercept represented each individual's TIV at their own baseline. This model was fit only to the 38 syndromic CLN3 participants; the model-derived baseline TIVs were then held constant across follow-up visits, producing an "adjusted TIV" specific to each individual's own natural history (eFigure 3B). Because vision-only participants already showed flat TIV trajectories, their raw BPVs were instead normalized by their own average TIV across visits. Since the early-childhood participants show developing characteristics (brain and the cranium still increasing with age – i.e., TIV not affected by pathology), the raw BPVs were simply divided by their corresponding TIVs at each visit.

The normalized proportion (raw BPV / adjusted TIVs) addressed two problems: variability from differences in head size and sex, and underestimation of atrophy. However, this ratio still misrepresented true parenchyma volume. Looking at eFigure 3C(i), participants near age 6 showed a relative BPV below 90%, implying over 10% atrophy at an age when little/no atrophy is expected. This reflects the fact that, at a healthy baseline, parenchyma occupies only ~87%–90% of the cranial cavity.[14] We therefore applied a further normalization step, scaling by the expected brain parenchyma fraction (BPF) in healthy adolescents and young adults.

We estimated the healthy BPF from three independent sources. A published meta-analysis restricted to healthy adults aged 21–35 years yielded a median BPF of 0.867.[14] We additionally computed BPF using our own pipeline in two independent pediatric datasets: 121 healthy participants aged 9–11 years (49% female) from the Adolescent Brain Cognitive Development (ABCD) Study,[15] and 38 healthy participants aged 9.5–11.3 years (42% female) from the Neuroimaging of the Children's Attention Project (NICAP).[16] Median BPF was 0.867 in the ABCD sample and 0.876 in NICAP. Averaging across all three sources yielded a BPF of 0.87, indicating that brain parenchyma comprises ~87% of intracranial volume from childhood through early adulthood in healthy individuals.

Using this healthy BPF, normalized BPV was computed as shown in equation [2] below:

$$Normalized\ BPV_{i,k} = \frac{\left(\frac{Raw\ BPV_{i,k}}{Adjusted\ TIV_{i,k}}\right)}{BPF_{Healthy}} \times 100\ (\%)$$

which simplifies to:

$$Normalized\ BPV_{i,k} = \frac{Raw\ BPV_{i,k}}{Adjusted\ TIV_{i,k}\ \times 0.87} \times 100\ (\%) \qquad [2]$$

where $BPF_{Healthy} = 0.87$. This fully normalized BPV appropriately centered around ~100% at age 6 (eFigure 3C(ii)) and declined progressively with age, reflecting an unbiased estimate of atrophy rate.

Normalization substantially reduced inter-individual variability attributable to head size (compare eFigures 3A(i) and 3C(ii)) while preserving the low intra-individual variability of raw BPV, supporting its reliability as a longitudinal measure. Throughout the remainder of this manuscript, "BPV"/"Normalized BPV" refer to the fully normalized values shown in eFigure 3C(ii).

The same approach was applied to each tissue compartment using compartment-specific healthy volume fractions derived from the same two pediatric databases. For example, the healthy GM volume fraction (GMVF) was 0.544 and 0.548 in the two databases, averaging to 0.55, giving equation [3]:

$$Normalized\ GMV_{i,k} = \frac{Raw\ GMV_{i,k}}{Adjusted\ TIV_{i,k}\ \times 0.55}\ \times 100\ (\%) \qquad [3]$$

The same process was repeated for WMV (Average WMVF = 0.32) and TCSFV (Average TCSFVF = 0.13) and its constituents PCSFV and VCSFV (Average PCSFVF = 0.12; Average VCSFVF = 0.0073, respectively).

### 2.0 eStatistical Analysis Plan (eSAP)

#### 2.1 Linear Mixed-Effect (LME) Model Framework

For each volumetric and clinical measurement, individual-level longitudinal trends were estimated using a linear mixed-effects (LME) model, with baseline age centered at 5 years (*Baseline_Age_C5*)– an approximate presymptomatic age to preserve a meaningful model intercept– as the between-participant predictor and longitudinal duration (*Individual_Duration*, an individual's longitudinal age centered by their own baseline age), as the within-participant predictor. The general model equation [4] is shown below with BPV as an example:

$$BPV_{ik} = \boldsymbol{\beta_0} + \boldsymbol{\beta_1}(Baseline\_Age\_C5_i) + \boldsymbol{\beta_2}(Individual\_Duration_{ik}) + \boldsymbol{u_{0i}} + \boldsymbol{u_{1i}}(Individual\_Duration_{ik}) + \boldsymbol{\varepsilon_{ik}} \qquad [4]$$

where, $\boldsymbol{\beta_0}$ is the population-average BPV at the reference baseline age of 5 years; $\boldsymbol{\beta_1}$ is the association between an individual's baseline age and their baseline BPV; $\boldsymbol{\beta_2}$ is the fixed-effect of interest representing the overall average longitudinal slope, $\boldsymbol{u_{0i}}$ and $\boldsymbol{u_{1i}}$ are participant-level random intercept and slope terms, and $\boldsymbol{\varepsilon_{ik}}$is residual error, for participant *i* at visit *k*.

For the Vineland Growth Scale Values (GSVs – Expressive, Personal, and Coping), a significant interaction between baseline age and longitudinal duration was observed; for these three measurements only, the model was extended to include this interaction term in equation [5], with Vineland Expressive GSV (VEGSV) as an example:

$$VEGSV_{ik} = \boldsymbol{\beta_0} + \boldsymbol{\beta_1}(Baseline\_Age\_C5_i) + \boldsymbol{\beta_2}(Individual\_Duration_{ik}) + \boldsymbol{\beta_3}(Baseline\_Age\_C5_i \times Individual\_Duration_{ik}) + \boldsymbol{u_{0i}} + \boldsymbol{u_{1i}}(Individual\_Duration_{ik}) + \boldsymbol{\varepsilon_{ik}} \quad [5]$$

For these subdomains, the average longitudinal slope was computed accounting for this interaction term, rather than from $\boldsymbol{\beta_2}$ alone.

All 95% confidence intervals reported in this study– including for LME fixed-effect estimates and $T_{min}$ (see below)– were computed via bootstrap resampling (N=2000 iterations).

### 2.2 Relative Standard Error (RSE)

To quantify the precision of each participant's individual-level slope relative to its magnitude, we computed a standard RSE for participant *i* as:

$$RSE_{Standard,i} = \frac{SE\_Slope_i}{|Slope_i|} \times 100 \quad [6]$$

where $SE\_Slope_i$ is the LME-derived standard error of participant *i*'s (random-effect) slope estimate. Lower RSE indicates greater precision, i.e., less uncertainty in the estimated slope relative to its magnitude. Because $RSE_{Standard,i}$ approaches infinity as a stable individual's slope approaches zero, we also computed a $RSE_{Normalized,i}$, in which each participant's $SE\_Slope_i$ was instead normalized by the magnitude of the overall average longitudinal slope (fixed-effect for $Individual\_Duration_{ik}$ in eq.[4]):

$$RSE_{Normalized,i} = \frac{SE\_Slope_i}{|Average\_Longitudinal\_Slope|} \times 100 \quad [7]$$

The $RSE_{Normalized,i}$ from eq.[7] is less sensitive to extreme values from near-zero individual slopes and was used alongside $RSE_{Standard,i}$ to characterize precision across the cohort.

### 2.3 Minimum Inter-Visit Interval ($T_{min}$)

$T_{min}$ was derived from the concept of minimal detectable change: the smallest true change in a measurement distinguishable from measurement noise. For participant $i$, with LME-estimated individual slope: $Slope_i$, let $Measurement\_Error_i$ denote the within-participant residual error– the typical deviation between observed data and the model-predicted values. Unlike the standard error of the slope, which scales with elapsed time, $Measurement\_Error_i$ is fixed for each participant's fitted trajectory. The accumulated true change after elapsed time $t$ is $Slope_i \times t$, which grows linearly with $t$; because the noise term does not, a finite elapsed time exists at which accumulated change first exceeds this threshold. For a one-sided (e.g., in the direction of disease-worsening/progression) 95% threshold: $Slope_i \times t = 1.645 \times Measurement\_Error_i$. Solving for $t$ gives the minimum elapsed time for participant $i$ to show a significant, directionally consistent change:

$$T_{min,i} = \frac{1.645 \times Measurement\_Error_i}{|Slope_i|} \qquad [8]$$

Participants with steeper slopes or lower measurement error reach significance sooner; those with smaller slopes or noisier measurements require longer follow-up. The empirical cumulative distribution of $T_{min}$ across participants was then used to identify the minimum inter-visit interval required for a specified proportion of the cohort (e.g., 75%: $T_{min,75\%}$)– to reach significant, directionally consistent change to reliably document CLN3 progression.

### eFigures

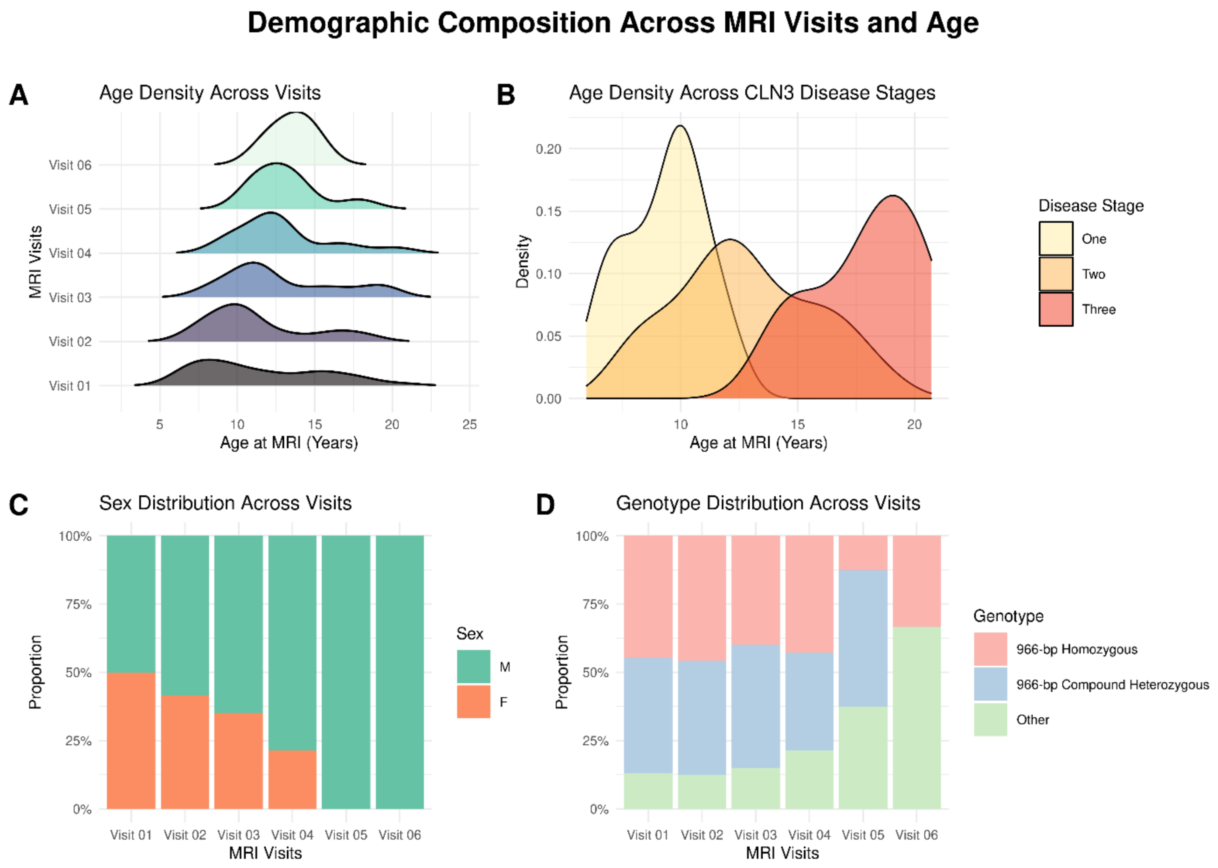


**eFigure 1. Participant demographic composition graphs for the MRI data for syndromic CLN3 participants.** The top two panels show the age density plots across MRI visits (**A**) and disease stages (**B**). Note the collinearity of the disease stage with age, e.g., Stage Three (light-red) peak appears much later than the peaks of Stages One and Two. The bottom two panels show the distribution in percentages by sex (**C**) and the genotype (**D**).

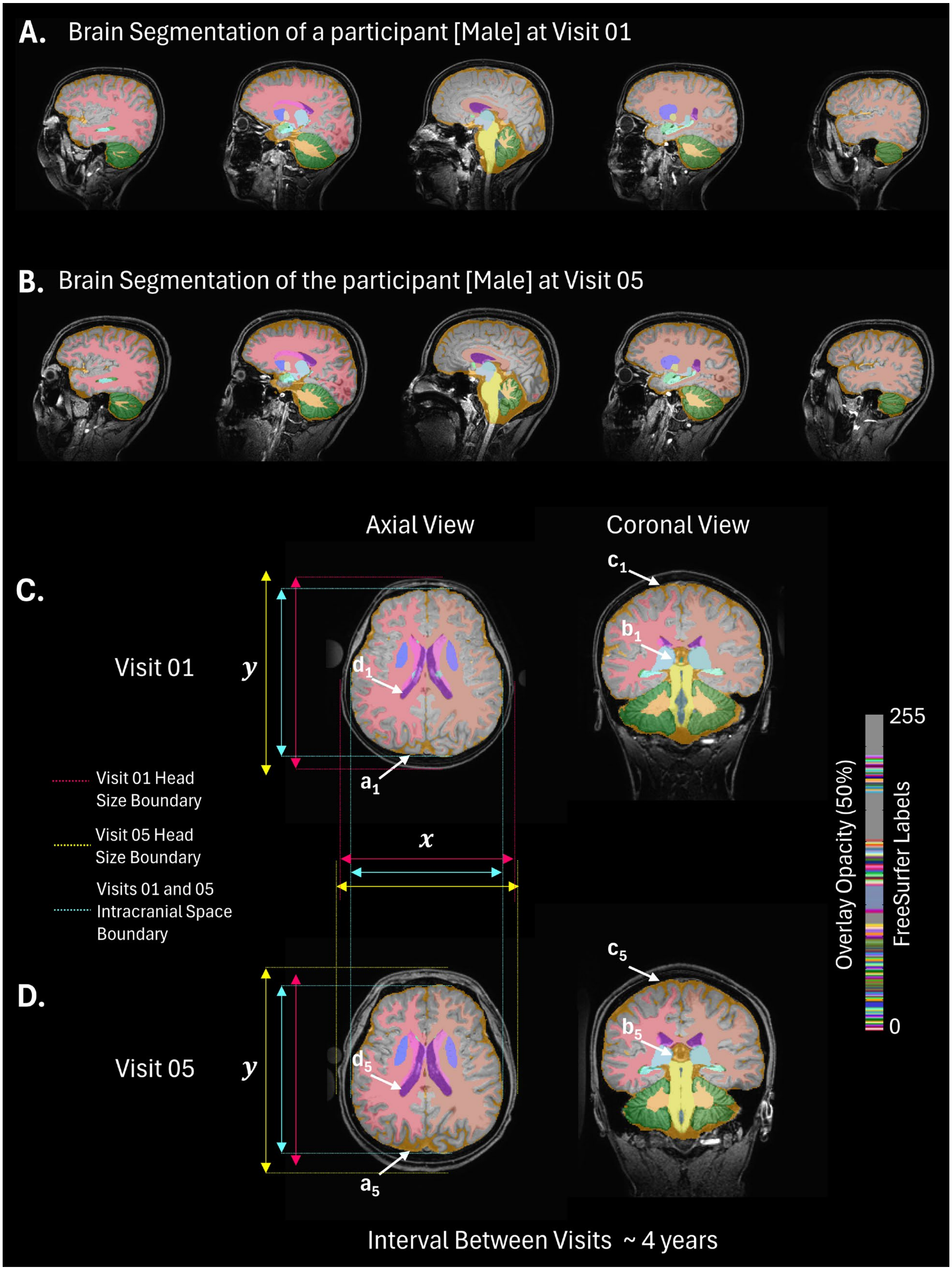
A. Brain Segmentation of a participant [Male] at Visit 01
B. Brain Segmentation of the participant [Male] at Visit 05
Axial View
Coronal View
C.
Visit 01
y
d1
a1
c1
b1
Visit 01 Head Size Boundary
Visit 05 Head Size Boundary
Visits 01 and 05 Intracranial Space Boundary
x
D.
Visit 05
y
d5
a5
c5
b5
255
0
Overlay Opacity (50%)
FreeSurfer Labels
Interval Between Visits ~ 4 years

**eFigure 2. Segmentation of T1w images using 'SynthSeg' and demonstrating parenchymal atrophy. A.** Segmentation of brain tissues in a CLN3 participant at the first MRI visit (Visit 01). A sagittal montage is shown that traverses the brain from the left-to-right ($x$ direction). **B.** Brain segmentation maps from the same participant, at the fifth visit (Visit 05). The total interval elapsed between Visits 01 and 05 is ~ 4 years. For better visual clarity and comparison, single slices from the axial and coronal planes are also shown for the same two visits in **C** and **D**, respectively. Looking at the axial slices in **C** and **D** – the head size increases between the first and fifth visits as the subject grows through ~ 4 years. The red edge lines and solid arrows show the extent of the head tissue boundaries at Visit 01, while the yellow edge lines and solid arrows show the extent at Visit 05. The difference in head size becomes clear from the lengths of the red and yellow arrows in both $x$ (horizontal) and $y$ (vertical) directions. The head-size boundary lengths (solid red and yellow) from both visits were placed side-by-side adjacent to each visit's axial image for easier comparison. The intracranial space boundaries remained virtually constant between the two visits (cyan). The subscript number in the label arrows (white) represents the visit at which the state of the pointed location is described. For example, $a_1$ and $a_5$ points to the peripheral CSF location at visits 01 and 05, respectively. The color-bar represents the same color codes defined for standard FreeSurfer segmentation labels. Note, the overlay opacity of the segmentation maps was reduced by 50% so that the underlying anatomy in the T1w images can be appreciated. The figure shows parenchymal atrophy and simultaneous increase in peripheral ($a_{1\text{-}5}$, $b_{1\text{-}5}$, $c_{1\text{-}5}$) and ventricular ($d_{1\text{-}5}$) CSF volumes between visits 01 and 05.

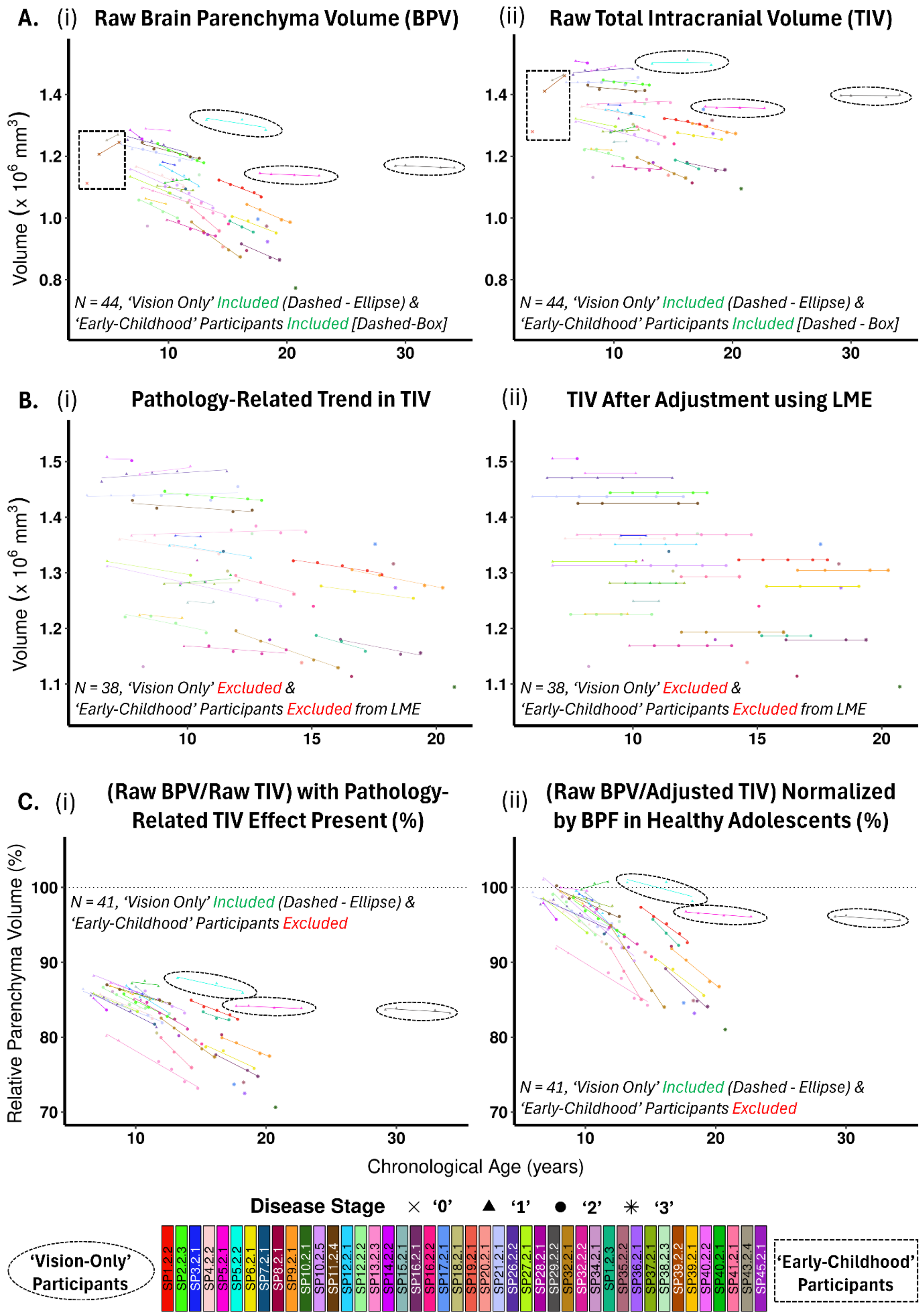
A. (i) Raw Brain Parenchyma Volume (BPV)
(ii) Raw Total Intracranial Volume (TIV)
Volume (x 10^6 mm^3)
N = 44, 'Vision Only' Included (Dashed - Ellipse) &
'Early-Childhood' Participants Included [Dashed-Box]
N = 44, 'Vision Only' Included (Dashed - Ellipse) &
'Early-Childhood' Participants Included [Dashed - Box]
B. (i) Pathology-Related Trend in TIV
(ii) TIV After Adjustment using LME
N = 38, 'Vision Only' Excluded &
'Early-Childhood' Participants Excluded from LME
N = 38, 'Vision Only' Excluded &
'Early-Childhood' Participants Excluded from LME
C. (i) (Raw BPV/Raw TIV) with Pathology-Related TIV Effect Present (%)
(ii) (Raw BPV/Adjusted TIV) Normalized by BPF in Healthy Adolescents (%)
Relative Parenchyma Volume (%)
N = 41, 'Vision Only' Included (Dashed - Ellipse) &
'Early-Childhood' Participants Excluded
N = 41, 'Vision Only' Included (Dashed - Ellipse) &
'Early-Childhood' Participants Excluded
Chronological Age (years)
Disease Stage '0' '1' '2' '3'
'Vision-Only' Participants
'Early-Childhood' Participants
SP1.2.2 SP2.2.3 SP3.2.1 SP4.2.2 SP5.2.1 SP5.2.2 SP6.2.1 SP7.2.1 SP8.2.1 SP9.2.1 SP10.2.1 SP10.2.5 SP11.2.4 SP12.2.1 SP12.2.2 SP13.2.3 SP14.2.2 SP15.2.1 SP16.2.1 SP16.2.2 SP17.2.1 SP18.2.1 SP19.2.1 SP20.2.1 SP21.2.1 SP26.2.2 SP27.2.1 SP28.2.1 SP29.2.2 SP32.2.1 SP32.2.2 SP34.2.1 SP1.2.3 SP35.2.2 SP36.2.1 SP37.2.1 SP38.2.3 SP39.2.2 SP39.2.1 SP40.2.2 SP40.2.1 SP41.2.1 SP43.2.4 SP45.2.1

**eFigure 3. Demonstrating the CLN3 pathology-related effect on total intracranial volume (TIV) measurements by SynthSeg and the process of normalizing the brain parenchyma volume (BPV) to measure atrophy.** The panels in each row share the y axes titles. The x-axis title (chronological age) is the same for all panels and shown commonly at the bottom. The axes limits were kept the same between the left and right panels in respective rows, for clarity of interpretation and comparison purposes. Each participant is represented by a unique color shown in the color-bar at the bottom. The 'Vision-Only' participants are shown with dashed ellipses and 'Early-Childhood' participants (ages 0-5 years) that show typical development are shown within dashed boxes (where applicable). The shape of data points represents the disease stage of an individual across their natural history. **A**(i) The raw brain parenchyma volumes (BPVs) computed from 'SynthSeg' segmentation using the T1w native space data. **A**(ii) The corresponding raw total intracranial volumes (TIVs) computed by 'SynthSeg' for each individual across MRI sessions. Both measurements show a decline with age at the individual level, but they are markedly higher (steeper slopes) for the raw BPVs, thereby indicating brain atrophy. Note, the 'Vision-Only' subjects (ellipse) did not show the same level of atrophy in BPVs as the syndromic CLN3 cohort (**A**(i)) and had nearly stable raw TIVs (**A**(ii)). **B**(i) The linear mixed effect (LME) model predicted fit of raw TIVs with age, at the individual level. Note, the 'Vision-Only' and the 'Early-Childhood' participants were removed for this analysis as they did not conform to the CLN3 cohort with classic presentation. **B**(ii) The LME-adjusted TIVs that reflect the best estimate of TIV at the baseline age of each individual. **C**(i) The BPVs after normalizing each individual's raw BPVs by their raw TIVs (essentially, **A**(i) divided by **A**(ii) times 100). Note, this underestimates the atrophy due to the pathology-related trends in the TIV measurements. **C**(ii) The correct representation of atrophy after normalizing raw BPVs by the

LME-adjusted TIVs at the individual level and further scaling this proportion with the best estimate of brain parenchyma fraction (BPF) in healthy adolescents (BPF = ~0.87). *Keys:* BPV = Brain Parenchyma Volume, TIV = Total Intracranial Volume, BPF = Brain Parenchyma Fraction

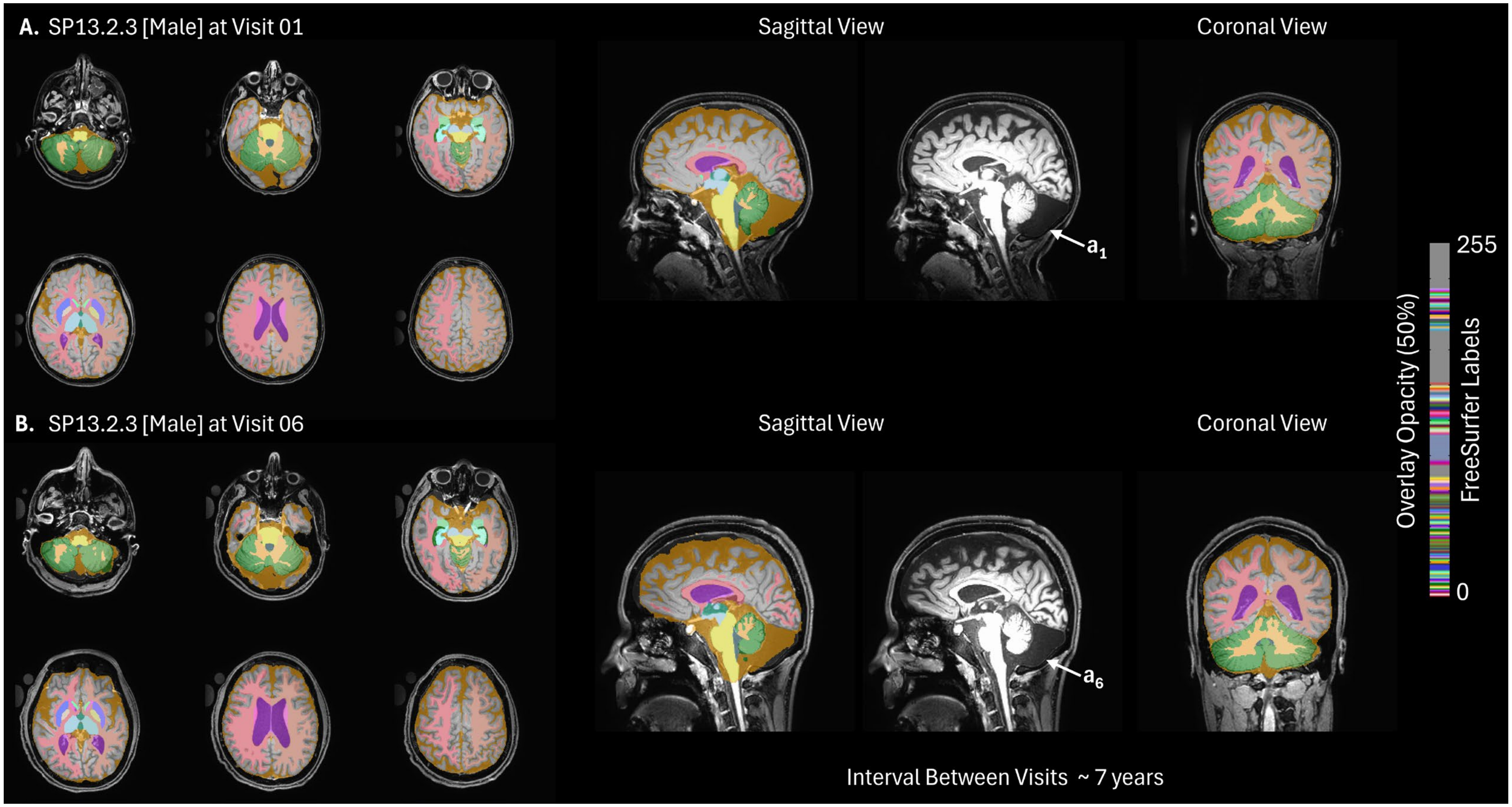


**eFigure 4. Brain segmentation of SP13.2.3 (male), with the arachnoid cyst.** The participant has had six MRI visits and segmentations from only visits 01 (**A**) and 06 (**B**) are shown to show the difference between the baseline and the last visit. The axial montage shows remarkable increase in the ventricular CSF (in purple) and peripheral CSF (in golden orange) volumes at visit 06 (**B**). Labels $a_1 a_6$ in the sagittal view shows the presence of a congenital arachnoid cyst in this participant at visits 01 and 06, respectively. Note that the size of the cyst remains stable throughout the six MRI visits. Furthermore, SynthSeg misclassified the CSF-like tissue

inside the cyst as part of the peripheral CSF – leading to overestimating the TIV. The figure shows that neurodegeneration (as is characteristic of CLN3) has progressed over the years, while the cyst remained approximately unchanged.

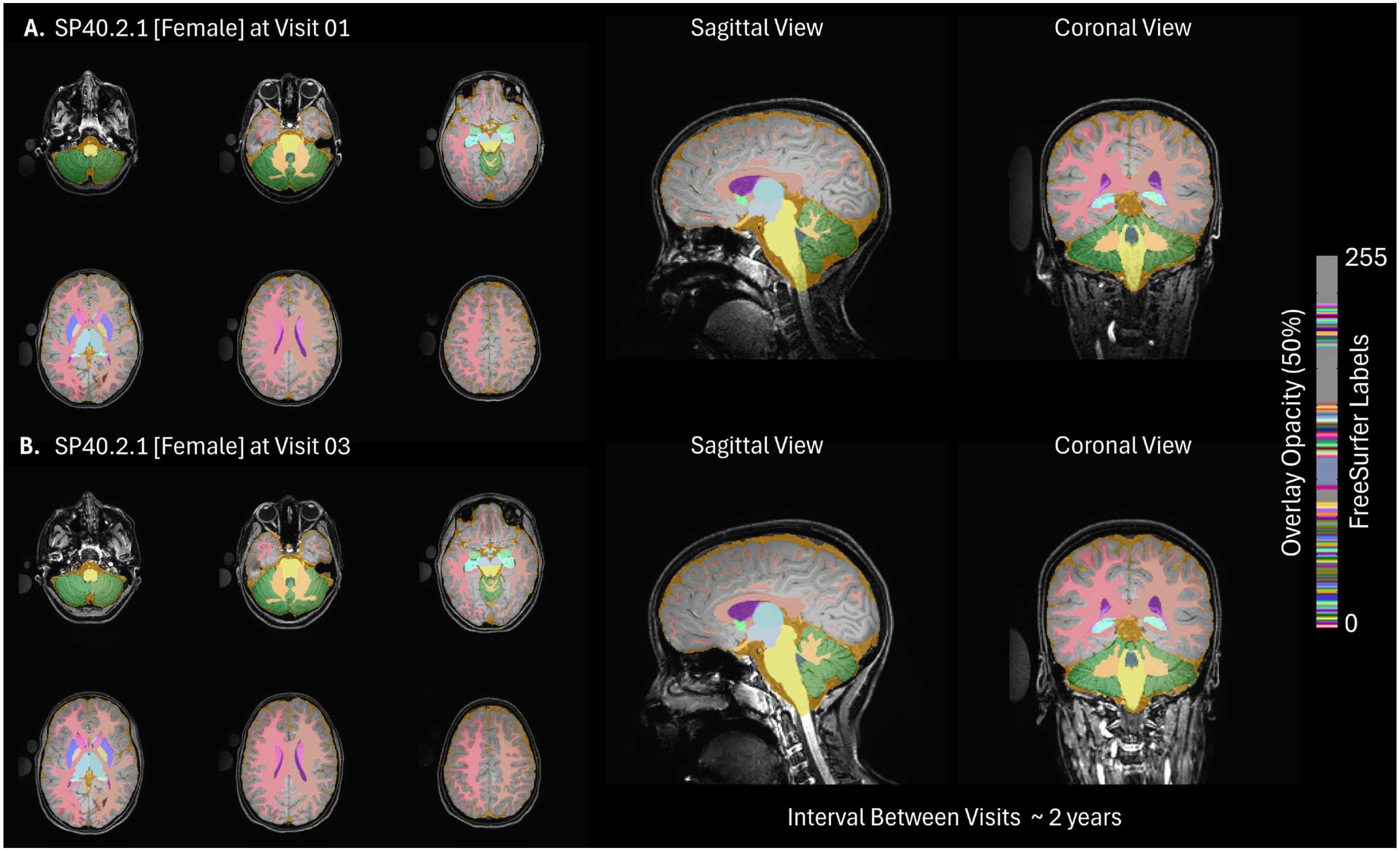


**eFigure 5. Brain Segmentation of SP40.2.1 (female), with relatively stable BPV across age.** This participant has had only three visits so far, with approximately one year gap between visits. Note that the brain looks fairly normal between the visits, resembling typically developing children in that age range. Also, note no remarkable signs of atrophy was observed between the visits.

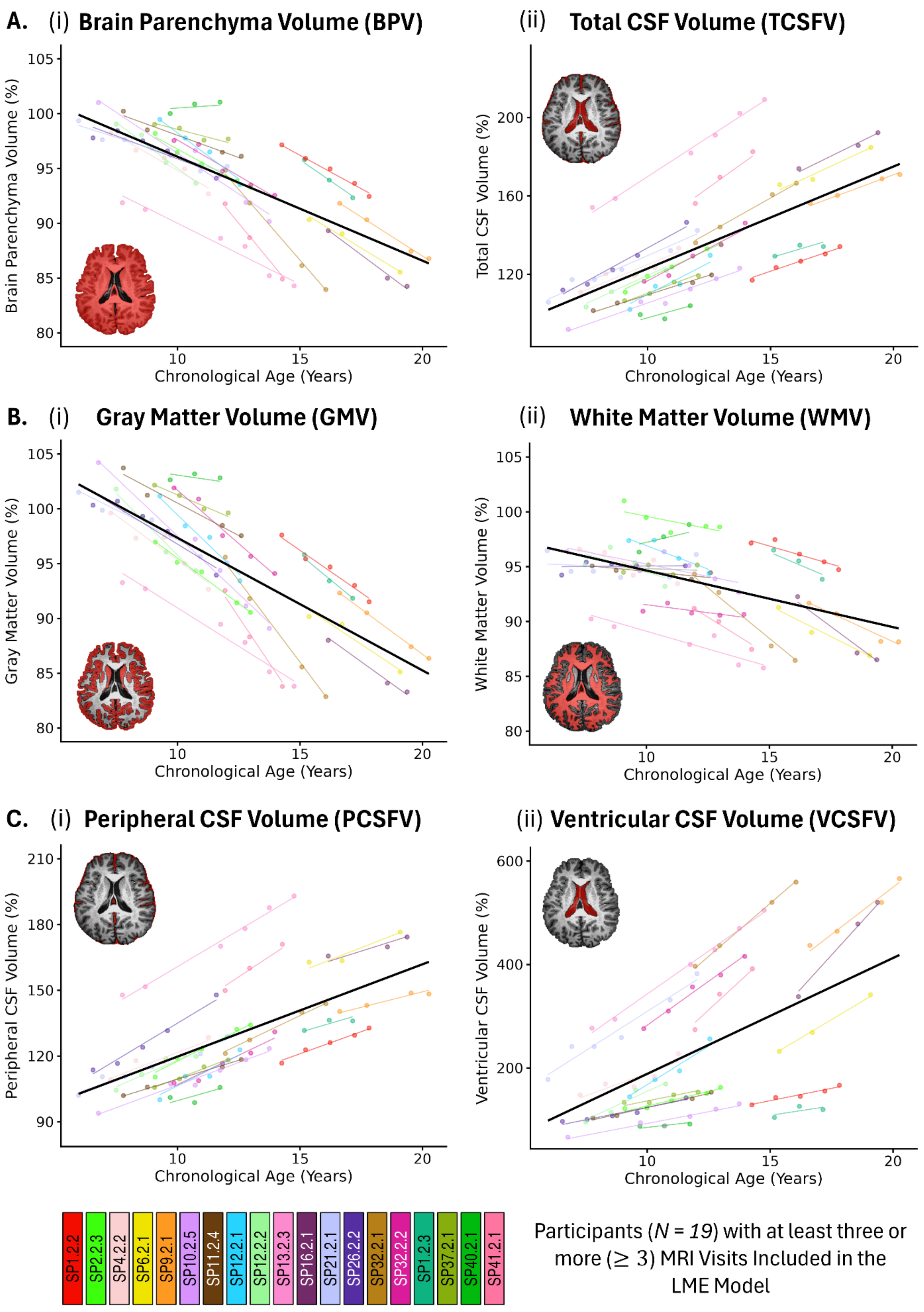


Participants ($N$ = 19) with at least three or more (≥ 3) MRI Visits Included in the LME Model

**eFigure 6. Linear Mixed-Effect (LME) model fitted individual and population trends for volumetric measurements.** A representative binary mask for each tissue class is provided in each panel for the corresponding brain compartment. The brain parenchyma (**A**(i)) consists of the combined gray and white matter tissues and therefore the red voxels cover more spatial extent. The rest of the space (excluding parenchyma) in the cranial cavity is filled by CSF (**A**(ii)). The BPV and GMV (**A**(i) and **B**(i), respectively) show strong decline in volumes with age at the individual and population level. The WMV (**B**(ii)) also shows decrease in volume but the effect is noticeably smaller compared to the GMV loss. The decrease in parenchyma volume must be compensated by increasing CSF volumes (**A**(ii)) to satisfy the Monro-Kellie doctrine. Both the peripheral and ventricular CSF (**C**(i) and **C**(ii), respectively) compartments reflect this increase in total CSF volume with age – confirming this biological process. In effect, the overall slopes from all measurements strongly point to disease progression, i.e., progressive brain atrophy. Note that all volumes are relative. Each tissue compartment volume is represented as percentage (%) of the expected volume fraction of that specific tissue compartment in healthy adolescent brains. Therefore, if there is atrophy (e.g., GMV and BPV), the volumes will naturally tend to go below 100% longitudinally, and on the contrary, if a tissue compartment is increasing in volume to offset the atrophy (e.g., TCSFV), the volumes will tend to go higher than 100% longitudinally.

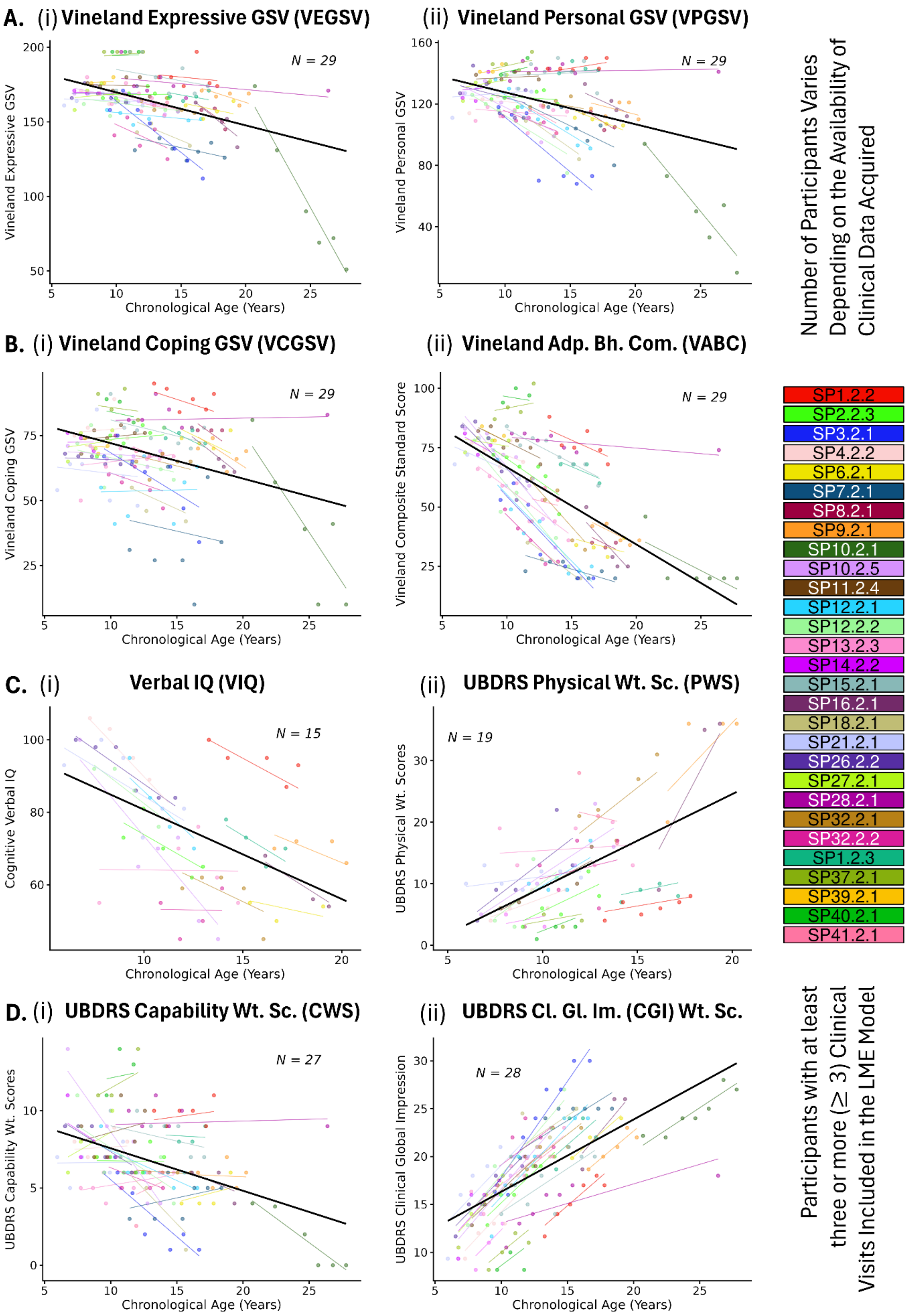
A. (i) Vineland Expressive GSV (VEGSV)
Vineland Expressive GSV
N = 29
Chronological Age (Years)
(ii) Vineland Personal GSV (VPGSV)
Vineland Personal GSV
N = 29
Chronological Age (Years)
B. (i) Vineland Coping GSV (VCGSV)
Vineland Coping GSV
N = 29
Chronological Age (Years)
(ii) Vineland Adp. Bh. Com. (VABC)
Vineland Composite Standard Score
N = 29
Chronological Age (Years)
C. (i) Verbal IQ (VIQ)
Cognitive Verbal IQ
N = 15
Chronological Age (Years)
(ii) UBDRS Physical Wt. Sc. (PWS)
UBDRS Physical Wt. Scores
N = 19
Chronological Age (Years)
D. (i) UBDRS Capability Wt. Sc. (CWS)
UBDRS Capability Wt. Scores
N = 27
Chronological Age (Years)
(ii) UBDRS Cl. Gl. Im. (CGI) Wt. Sc.
UBDRS Clinical Global Impression
N = 28
Chronological Age (Years)
Number of Participants Varies Depending on the Availability of Clinical Data Acquired
SP1.2.2
SP2.2.3
SP3.2.1
SP4.2.2
SP6.2.1
SP7.2.1
SP8.2.1
SP9.2.1
SP10.2.1
SP10.2.5
SP11.2.4
SP12.2.1
SP12.2.2
SP13.2.3
SP14.2.2
SP15.2.1
SP16.2.1
SP18.2.1
SP21.2.1
SP26.2.2
SP27.2.1
SP28.2.1
SP32.2.1
SP32.2.2
SP1.2.3
SP37.2.1
SP39.2.1
SP40.2.1
SP41.2.1
Participants with at least three or more (≥ 3) Clinical Visits Included in the LME Model

**eFigure 7. Linear Mixed-Effect (LME) model fitted individual and population trends for clinical measurements.** All Vineland Growth Scale Values (GSVs) (**A**(i)-(ii), **B**(i)-(ii)) show an overall decrease with age; with the VABC (**B**(ii)) showing a stronger and more consistent decline at the individual level. Similarly, verbal IQ (**C**(i)) shows a strong decline with age. The UBDRS physical and capability weighted scores (PWS: **C**(ii) and CWS: **D**(i)) show an overall increase and decrease with age, respectively). The UBDRS clinical global impression (CGI: **D**(ii)) weighted scores  show a strong positive increase suggesting disease progression. To summarize, all clinical measurements, except for UBDRS CWS, showed a statistically significant overall longitudinal effect with age that aligns with disease progression, although some variability in longitudinal trends can be observed across individuals. *Keys:* GSV = Growth Scale Values; Vineland Adp.. Bh. Com. = Vineland Adaptive Behavior Composite (VABC); Wt. Sc. = Weighted Scores, UBDRS Cl. Gl. Im. = Unified Batten Disease Rating Scale Clinical Global Impression

8. Kellie G. An Account of the Appearances Observed in the Dissection of Two of Three Individuals Presumed to Have Perished in the Storm of the 3d, and Whose Bodies Were

Discovered in the Vicinity of Leith on the Morning of the 4th, November 1821; with Some Reflections on the Pathology of the Brain: Part I. Trans Med Chir Soc Edinb 1824;1:84-122.

9. Monro A. Observations on the structure and functions of the nervous system: Classics of Neurology & Neurosurgery Library Division of Gryphon Editions, 1994.
